# Age- and Sex-specific Reference Ranges for Cardiac Function and Structure in Germany: Cardiovascular Magnetic Resonance Imaging (CMR) in the German National Cohort (NAKO)

**DOI:** 10.64898/2026.03.09.26347892

**Authors:** Christopher L. Schlett, Christopher Schuppert, Peter M. Full, Robin T. Schirrmeister, Manuel Hein, Marco Reisert, Maximilian F. Russe, Martyna Flis, Jan Gröschel, Clemens Ammann, Valeria Geiger, Karin Halina Greiser, Tafirenyika Gwenzi, Anna Köttgen, Thomas Kröncke, Thomas Küstner, Wolfgang Lieb, Lea Jigme Michel, Konstantin Nikolaou, Annette Peters, Tobias Pischon, Henning Teismann, Henry Völzke, Klaus H. Maier-Hein, Fabian Bamberg, Susanne Rospleszcz, Jeanette Schulz-Menger

**Affiliations:** Department of Diagnostic and Interventional Radiology, Medical Center - University of Freiburg, Faculty of Medicine, University of Freiburg, Freiburg, Germany; Division of Medical Image Computing, German Cancer Research Center (DKFZ), Heidelberg, Germany; Medical Faculty Heidelberg, Heidelberg University, Heidelberg, Germany; Medical Physics, Department of Diagnostic and Interventional Radiology, Medical Center - University of Freiburg, Faculty of Medicine, University of Freiburg, Freiburg, Germany; Department of Cardiology and Angiology, University Heart Center Freiburg - Bad Krozingen, Medical Center - University of Freiburg, Faculty of Medicine, University of Freiburg, Freiburg, Germany; Department of Stereotactic and Functional Neurosurgery, Medical Center - University of Freiburg, Faculty of Medicine, University of Freiburg, Freiburg, Germany; Charité - Universitätsmedizin Berlin, corporate member of Freie Universität Berlin and Humboldt Universität zu Berlin, Berlin, Germany; Working Group on CMR, Experimental and Clinical Research Center, Max Delbrück Center for Molecular Medicine in the Helmholtz Association and Charité - Universitätsmedizin Berlin, Berlin, Germany; Deutsches Herzzentrum der Charité, Department of Cardiology, Angiology and Intensive Care Medicine, Charitéplatz 1, 10117 Berlin, Germany; DZHK (German Centre for Cardiovascular Research), partner site Berlin, Berlin, Germany; Division of Cancer Epidemiology, German Cancer Research Center (DKFZ) Heidelberg, Heidelberg, Germany; Division of Clinical Epidemiology of Early Cancer Detection, German Cancer Research Center (DKFZ), Heidelberg, Germany; Institute of Epidemiology and Prevention, Medical Center-University of Freiburg, Faculty of Medicine, University of Freiburg, Freiburg, Germany; Department of Diagnostic and Interventional Radiology, University Hospital Augsburg, Augsburg, Germany; Centre for Advanced Analytics and Predictive Sciences, University of Augsburg, Augsburg, Germany; Department of Diagnostic and Interventional Radiology, University Hospital Tuebingen, Tuebingen,Germany; Institute of Epidemiology, Kiel University, Kiel, Germany; Institute of Epidemiology, Helmholtz Zentrum München - German Research Center for Environmental Health (GmbH), Neuherberg, Germany; Chair of Epidemiology, Institute for Medical Information Processing, Biometry and Epidemiology, Medical Faculty, Ludwig-Maximilians-Universität München, Munich, Germany; DZHK (German Centre for Cardiovascular Research), partner site Munich Heart Alliance, Munich, Germany; Max Delbrück Center for Molecular Medicine in the Helmholtz Association (MDC), Molecular Epidemiology Research Group, Berlin, Germany; Max Delbrück Center for Molecular Medicine in the Helmholtz Association (MDC), Biobank Technology Platform, Berlin, Germany; Institute of Epidemiology and Social Medicine, University of Münster, Münster, Germany; Institute for Community Medicine, University Medicine Greifswald, Germany; Department of Radiation Oncology, Medical Faculty, Heidelberg University, Heidelberg, Germany

## Abstract

**Introduction:** Cardiovascular magnetic resonance (CMR) is the reference standard for quantifying cardiac structure and function, yet widely applicable population-based reference values remain limited. We derived age- and sex-specific reference ranges for ventricular volumes, mass, and function using data from the population-based German National Cohort (NAKO).

**Methods:** Short-axis balanced steady-state free precession cine images from 3T CMR of 29,908 participants were analyzed using a validated deep learning segmentation pipeline with systematic quality control. From these, we defined a main reference cohort free of cardiovascular disease (CVD), and a healthy subcohort additionally free of cardiovascular risk factors. Standard left (LV) and right ventricular (RV) measures were quantified and indexed. Reference intervals (5th–95th percentiles) were modeled using additive models and quantile regression to capture non-linear age trends, stratified by sex, with formal testing for age-sex interactions.

**Results:** The CVD-free reference cohort included 24,371 participants (mean age 43.8±12.2 years; age range 20-72 years, 44.7% women). LV and RV end-diastolic and end-systolic volumes declined with age, whereas LV ejection fraction remained stable and RV ejection fraction increased modestly. Sex differences were consistent across metrics and all major parameters demonstrated significant age-sex interactions; differences were most pronounced at younger ages and attenuated in later life. The healthy subcohort (n=5,550) showed similar structural and functional profiles, without clinically relevant deviations from the main reference cohort.

**Conclusions:** This study provides age- and sex-specific CMR reference ranges derived from a large, uniformly imaged national cohort. These data offer a population-based normative framework for clinical CMR interpretation and future research on sex-specific cardiac remodeling and healthy aging.

## Background

Cardiovascular imaging plays a crucial role in clinical decision-making. Measurements of cardiac chamber size and function derived from imaging are fundamental for cardiovascular risk stratification, diagnosis, and therapeutic planning. Since cardiovascular magnetic resonance (CMR) is considered to be the clinical reference standard for evaluating cardiac structure and function, the accurate differentiation between health and disease using this modality is essential to provide best clinical care to patients with cardiovascular health issues.

Substantial efforts have been undertaken to establish reliable reference ranges for CMR parameters, including carefully designed studies in healthy volunteers, multicenter registries, and large population-based cohorts. Early work focused on smaller, well-characterized cohorts of individuals without known cardiac disease, which laid the foundation for subsequent specific reference values. A recent meta-analysis reviewed 254 studies from 25 different countries, and provided CMR reference values for up to 4000 individuals free of cardiovascular disease (CVD) [1].

Additionally, large population-based analyses have been conducted, most notably within the UK Biobank (UKB). An early analysis identified and characterized CMR parameters of 804 healthy individuals among 5,065 participants [2]. A subsequent analysis performed within the framework of the Healthy Hearts Consortium - incorporating data from the UKB alongside several additional, predominantly smaller European cohorts - expanded this effort using more recent data and identified 7,672 healthy individuals out of 50,018 participants [3]. The relatively small proportion of healthy subjects in the UKB is largely attributable to the older age of participants at the time of CMR acquisition (45 - 74 years; [2]). Across the entire Healthy Hearts Consortium, the number of included individuals younger than 40 years did not exceed 500.

The German National Cohort (NAKO) is among the world’s largest population-based prospective studies, established to investigate the causes of widespread diseases, identify risk factors, and improve early detection and prevention strategies. In addition to the collection of extensive baseline questionnaire data, biological samples, and physical measurements, whole-body MR imaging - including standard CMR sequences - was performed in more than 30,000 participants across a broad age range [4]. Given the growing evidence that biological heart age closely correlates with chronological age [5], it is imperative that definitions of “normal” cardiac parameters strongly account for age-related differences. Beyond age, the fundamental question remains: how should cardiovascular health be defined beyond the mere absence of overt disease?

Based on the MR substudy of the NAKO, we established CMR reference values for left (LV) and right ventricular (RV) structure and function in a cohort free of CVD, as well as in a subcohort free of both CVD and cardiovascular risk factors. Reference values were further provided for age- and sex-specific subgroups and analyzed with respect to sex-related interactions across the age groups.

## Methods

### Study Design

The analysis is based on the NAKO study, a large, ongoing prospective cohort conducted across 18 regional examination centers by a network of 25 institutions in Germany. The primary aim of NAKO is to identify risk factors contributing to the development of major chronic diseases, including cancer, diabetes, cardiovascular, neurodegenerative, psychiatric, respiratory, and infectious conditions. Between 2014 and 2019, a total of 205,415 individuals from the general population, aged 19 to 74 years, were recruited during the baseline assessment. A nested MR imaging substudy was carried out at five specialized imaging centers, where 30,868 participants underwent whole-body MR imaging. Further details on the study design, study protocol and participant recruitment have been described elsewhere [4, 6, 7]. The NAKO Use and Access Committee approved this project based on the participants’ informed consent, its accordance with the aims of the NAKO, and ethical approval from the Ethics Committee of the Medical Faculty of Freiburg University, Germany (No. 20-1107).

### CMR Acquisition

MR imaging was conducted using identical 3 T whole-body scanners (MAGNETOM Skyra, Siemens Healthineers, Erlangen, Germany) operating with an identical software version across all five imaging sites. The imaging protocol included a cardiac assessment employing functional and quantitative techniques, notably the acquisition of a steady-state free precession (SSFP) full-cycle cine stack in the short-axis (SAX) orientation. This comprised 12 slices covering the heart from base to apex, with 25 phases evenly distributed throughout the cardiac cycle. Cardiac planes were automatically planned using vendor-provided software (Cardiac Dot Engine, Siemens Healthineers, Erlangen, Germany). A comprehensive technical description of the whole-body MR imaging procedures in NAKO has been published previously [8].

### CMR Image Analysis and Quality Control

Short-axis cine images were available for 29,908 of the 30,868 MR participants. A deep learning-based segmentation algorithm, previously developed and successfully validated in CMR image segmentation challenges, was subsequently applied [9]. This nnU-Net-based method integrates an ensemble of U-Net architectures: five operating in two dimensions, processing individual slices, and five operating in three dimensions, leveraging contextual information across slices. This ensemble enhances segmentation performance by compensating for each model’s individual weaknesses. This algorithm produced full-cycle segmentations of the LV endocardium and epicardium and the RV endocardium in 29,609 participants, while 299 cases could not be processed. Segmentation-derived measures included biventricular end-diastolic and end-systolic volumes (LVEDV, RVEDV and LVESV, RVESV), stroke volumes (LVSV, RVSV), and ejection fractions (LVEF, RVEF), as well as left-ventricular mass calculated at end-diastole using a myocardial density of 1.055 g/ml. End-diastolic myocardial wall thickness was assessed using a 16-segment model, and mid-septal wall thickness was defined as the average of segments 8 and 9. Papillary muscles were included in the blood pool and excluded from mass calculations [9, 10].

As part of data quality control, outliers were identified based on extreme values of segmentation-derived measures. In addition to the parameters listed above, these comprised the interventricular SV difference, interventricular and intraventricular phase differences, and abnormal LV time-volume curves. All short-axis cine images flagged as outliers underwent structured visual review, using predefined criteria to assess deficiencies affecting image quality (including artifact presence, alignment issues, slice shifts, missing slices) and segmentation accuracy (including oversegmentation, undersegmentation, label transposition errors). Each domain was rated independently on a five-point scale: 5-complete confidence (no relevant deficiencies), 4-high confidence (minor deficiencies unlikely to substantially affect derived parameters), 3-moderate confidence (deficiencies that may have a non-negligible parameter-dependent effect), 2-low confidence (major deficiencies resulting in substantially reduced reliability of derived parameters), 1-unusable (not suitable for reliable parameter extraction). A detailed description of the quality control framework, including all predefined classification criteria used for rating, has been published previously [9]. Only participants with ratings of complete (5) or high (4) confidence in both domains were included in the final analysis.

### Covariates

All participants underwent standardized examinations, interviews, as well as sampling of biospecimens at the study centers. The assessment of covariates has been described previously [6, 7]. Briefly, age was calculated based on self-reported date of birth and the date of the baseline examination. The MR scan took place after the baseline examination (median time 15 days, interquartile range 35 days). Sex was self-reported at baseline as male or female.

Weight and height were measured without shoes and in light clothing using a calibrated electronic scale (seca type 635, seca GmbH & Co. KG, Hamburg, Germany or equivalent), and a calibrated stadiometer (seca 274 or equivalent), respectively. BMI (body mass index) and BSA (body surface area, duBois formula) were derived from measurements of weight and height. Obesity was defined as a BMI of 30 kg/m² or higher.

Presence of CVD was defined by self-reported medical professional’s diagnosis of any of the following: myocardial infarction, narrowing of coronary arteries, heart failure, cardiac arrythmias, stroke, or peripheral artery disease.

Diabetes was defined by any of the following: self-reported physician diagnosis of diabetes, use of glucose-lowering medication, or HbA1c ≥6.5%. Hypercholesterolemia was defined by any of the following: Self-reported physician diagnosis of hypercholesterolemia, use of lipid-lowering medication, or LDL cholesterol ≥160 mg/dl. Hypertension was defined by any of the following: Self-reported physician diagnosis of hypertension, use of antihypertensive medication, or measured blood pressure ≥140/90 mmHg. Ever smoking was defined as current or former smoking, based on self-report.

CVD risk was estimated by the Framingham Risk Score for 10- and 30-year risk [11, 12], as well as the SCORE2 risk score [13].

### Statistical analysis

Two samples were defined: the main reference cohort, which excluded participants with known CVD, and a healthy subcohort, from which participants with traditional cardiovascular risk factors (diabetes, hypercholesterolemia, hypertension, obesity, and smoking as defined above) were additionally excluded (**Figure 1**).

**Figure 1:**
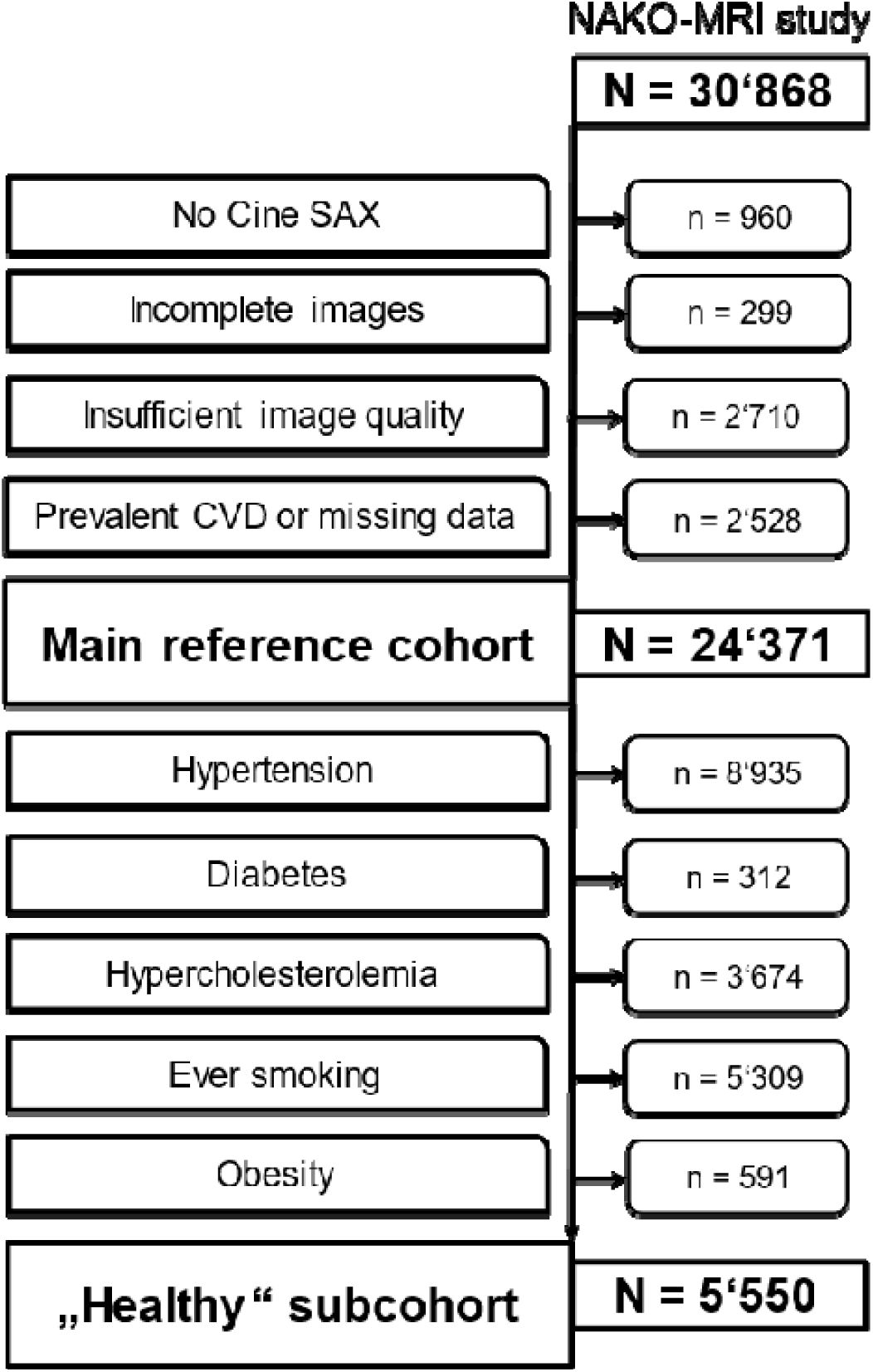
Participants’ flowchart

Participant characteristics and CMR metrics are described as means and standard deviations for continuous data, and counts and percentages for categorical data. Reference values for CMR metrics were derived by generalized additive models with a cubic spline basis and up to 20 knots, allowing for non-linear effects of age. The reference interval was derived by additive quantile regression, fitting the 5^th^ and 95^th^ percentile of distribution of the respective CMR metric, while allowing the conditional distribution of the CMR metric to vary with age.

Calculations were stratified by sex. Differences in the relationship between age and CMR metric according to sex were evaluated by visualization of differences in smoothing terms, and quantified by F-test between two additive models with an interaction term for age:sex. R^2^ values from a generalized additive model with and without CVD risk factors were compared.

R version 4.5.1 with libraries mcgv, qgam and gratia was used for analysis. A two-sided p-value <0.05 is considered to denote statistical significance.

## Results

### Study Cohorts

After excluding individuals with no CMR or with insufficient image quality (n=3’969) and prevalent CVD or unclear CVD history (n=2’528), the main reference cohort comprised 24’371 individuals (44.7% women, **Table 1**). The mean age was 47.6 years; the age range was 20 to 72 years while the majority was between 30 and 60 years old. The overall cardiovascular risk was low to moderate (median 10-yr Framingham risk: 8.0% [IQR: 3.4-16.3%], SCORE2: 2.6% [IQR: 1.1-5.2%]), and the prevalence of hypertension, hypercholesterolemia and diabetes was 36.7%, 31.8% and 4.4%, respectively (**Table 1**).

**Table 1:** Participant characteristics.

|  | Main reference cohort |  |  | Healthy subcohort |  |  |
| --- | --- | --- | --- | --- | --- | --- |
|  | All | Women | Men | All | Women | Men |
|  | N = 24'371 | N = 10'896<br>(44.7%) | N = 13'475<br>(55.3%) | N = 5'550 | N = 2'802<br>(50.5%) | N = 2'748<br>(49.5%) |
| Age, years | 47.6 ± 12.2 | 48.3 ± 12.1 | 47.0 ± 12.2 | 40.7 ± 12.0 | 41.8 ± 12.0 | 39.6 ± 11.8 |
| **MR imaging site** |  |  |  |  |  |  |
| Augsburg | 5'007 (20.5%) | 2'229 (20.5%) | 2'778 (20.6%) | 1'085 (19.5%) | 598 (21.3%) | 487 (17.7%) |
| Mannheim | 4'746 (19.5%) | 2'063 (18.9%) | 2'683 (19.9%) | 1'293 (23.3%) | 623 (22.2%) | 670 (24.4%) |
| Essen | 4'708 (19.3%) | 2'065 (19.0%) | 2'643 (19.6%) | 1'182 (21.3%) | 569 (20.3%) | 613 (22.3%) |
| Berlin | 4'722 (19.4%) | 2'034 (18.7%) | 2'688 (19.9%) | 1'140 (20.5%) | 532 (19.0%) | 608 (22.1%) |
| Neubrandenburg | 5'188 (21.3%) | 2'505 (23.0%) | 2'683 (19.9%) | 850 (15.3%) | 480 (17.1%) | 370 (13.5%) |
| **Anthropometry** |  |  |  |  |  |  |
| Height, cm | 173.0 ± 9.6 | 165.5 ± 6.6 | 179.1 ± 7.0 | 173.5 ± 9.6 | 166.8 ± 6.6 | 180.4 ± 7.0 |
| Weight, kg | 79.3 ± 16.1 | 70.9 ± 14.4 | 86.1 ± 14.0 | 71.8 ± 12.2 | 64.4 ± 8.8 | 79.4 ± 10.4 |
| BMI, kg/m<sup>2</sup> | 26.4 ± 4.6 | 25.9 ± 5.2 | 26.8 ± 4.0 | 23.7 ± 2.8 | 23.1 ± 2.9 | 24.4 ± 2.7 |
| BSA, m<sup>2</sup> | 1.93 ± 0.22 | 1.78 ± 0.17 | 2.05 ± 0.17 | 1.85 ± 0.19 | 1.72 ± 0.13 | 1.99 ± 0.15 |
| **Cardiovascular Risk** |  |  |  |  |  |  |
| SCORE2, % | 2.6 [1.1, 5.2] | 1.6 [0.7, 3.5] | 3.6 [1.8, 6.5] | 0.9 [0.4, 2.0] | 0.7 [0.2, 1.2] | 1.5 [0.7, 2.7] |
| FRS10, % | 8.0 [3.4, 16.3] | 5.1 [2.5, 10.1] | 11.6 [5.3, 21.5] | 3.0 [1.3, 6.0] | 2.5 [1.1, 4.1] | 4.5 [1.6, 8.3] |
| FRS30, % | 27.3 [13.9, 44.3] | 20.2 [10.6, 35.1] | 33.6 [18.8, 49.9] | 12.5 [5.8, 21.1] | 10.7 [4.4, 16.6] | 15.9 [7.0, 25.7] |
| Hypertension | 8'933 (36.7%) | 3'365 (30.9%) | 5'568 (41.3%) | 0* | 0* | 0* |
| Diabetes | 1'081 (4.4%) | 468 (4.3%) | 613 (4.5%) | 0* | 0* | 0* |
| Obesity | 4'657 (19.1%) | 2'077 (19.1%) | 2'580 (19.2%) | 0* | 0* | 0* |
| Hypercholesterolemia | 7'745 (31.8%) | 3'203 (29.4%) | 4'542 (33.7%) | 0* | 0* | 0* |
| Ever Smoking | 11'739 (48.2%) | 4'829 (44.3%) | 6'910 (51.3%) | 0* | 0* | 0* |

Values are presented as mean ± standard deviation, or median [Q1, Q3] for continuous variables, and counts with percentages for categorical variables. \*Not present due to definition of the healthy subcohort.
BMI: Body mass index; BSA: body surface area; FRS: Framingham Risk Score.

The healthy subcohort without cardiovascular risk factors (n=5,550; **Figure 1**; 23% of the main reference cohort) was younger and included more women compared to the main reference cohort (40.7 vs. 47.6 years and 50.5% vs. 44.7% women). As expected from the definition of this subcohort, the CVD risk scores yielded lower values (e.g. median 10-yr Framingham risk: 3.0% [IQR: 1.3-6.0%], SCORE2: 0.9 [IQR: 0.4-2.0%]). Notably, the MR imaging site Neubrandenburg exhibited the lowest inclusion rate for the healthy subcohort (16.4%, **Supplementary Table 1**), primarily attributable to the highest proportion of hypertension (46.1%) and obesity (24.0%) at this site (**Supplementary Table 1**). BMI and BSA were lower in the healthy subcohort (**Table 1**) due to the exclusion of individuals with obesity.

### CMR Metrics

In the main reference cohort, average LVEF was 63.4 % while LVEDV_bsa and LVESV_bsa were 73.8 ml/m² and 52.4 ml/m², respectively (**Supplementary Table 2**). Average LV Mass_bsa was 58.4 g/m² with a thickness of the septum of 7.8 mm. RVEF was 56.1 % while RVEDV_bsa and RVESV_bsa were 83.0 ml/m² and 36.6 ml/m², respectively. Cardiac Index (CO_bsa) was 3.0 l/min/m^2^ for LV and RV (**Supplementary Table 2**).

### Age- and Sex-specific Reference Ranges

The reference ranges, defined from the fitted 5^th^ to 95^th^ quantile, varied accordingly to the different CMR metrics, e.g. the reference ranges comprised about 16-18 percent-points for LVEF and about 80 ml for RVEDV (**Figure 2**, **Table 2a and 2b**).

**Figure 2:**
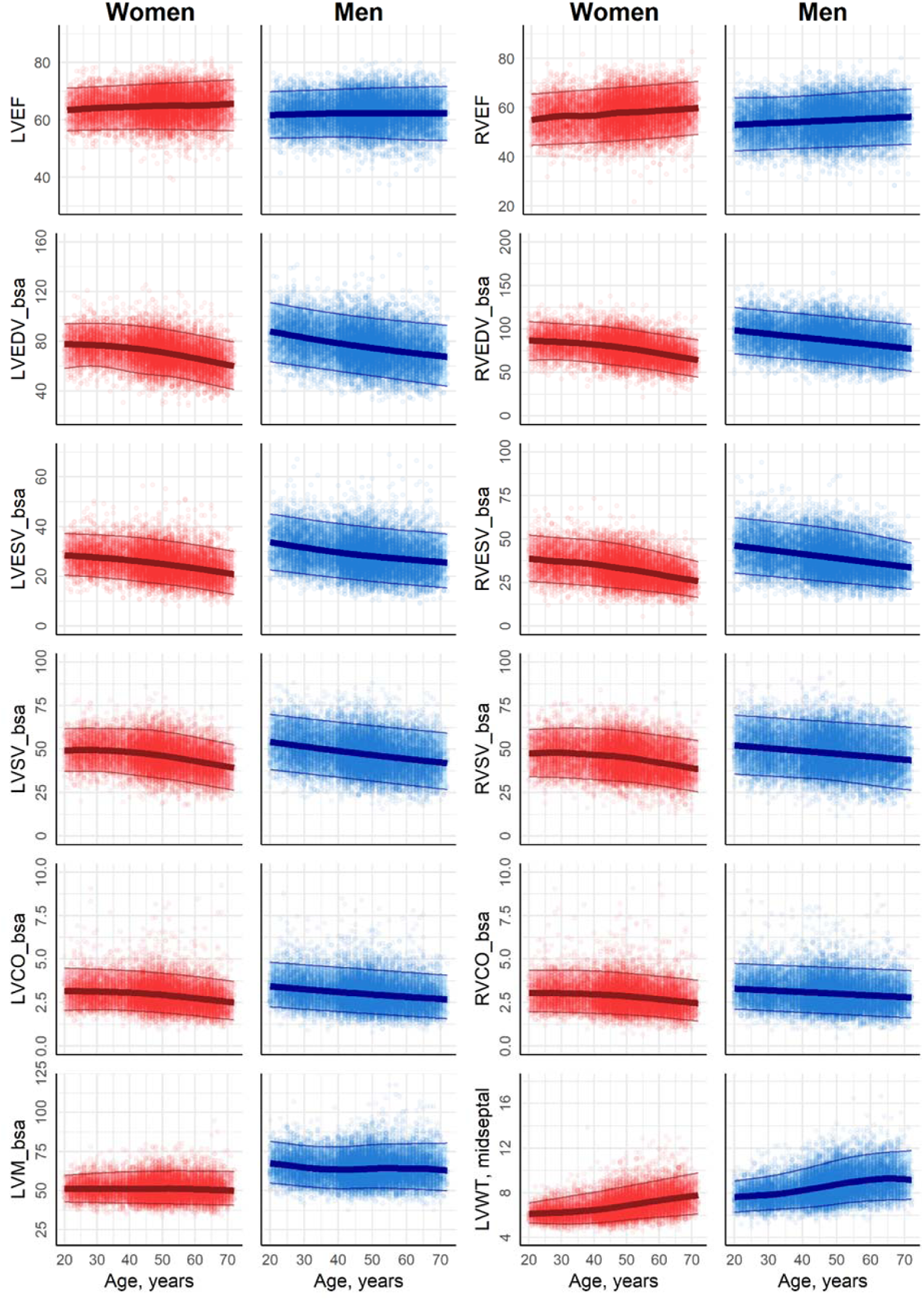
CMR reference ranges according to continuous age Figures show fitted mean (5th quantile, 95th quantile) of the CMR metric on the y-axis, and continuous age on the x-axis for women (red) and men (blue). LV: left ventricle; RV: right ventricle; EF: ejection fraction; EDV: end-diastolic volume; ESV: end-systolic volume; SV: stroke volume; CO: cardiac output; LVM: Left ventricular mass; WT: wall thickness; _bsa: indexed to body surface area. EF is given in %. EDV_bsa, ESV_bsa, and SV_bsa are given in ml/m^2^. LVCO_bsa is given in l/min/m^2^. LVM_bsa is given in g/m^2^. LVWT is given in mm.

**Table 2a:** CMR reference ranges for women.

| Age group |  |  |  |  |  |  |  |  |  |  |
| --- | --- | --- | --- | --- | --- | --- | --- | --- | --- | --- |
| years | [20,25) | [25,30) | [30,35) | [35,40) | [40,45) | [45,50) | [50,55) | [55,60) | [60,65) | [65,72] |
| N (% of main reference cohort) | 517 (3.8%) | 1'032 (7.7%) | 986 (7.3%) | 877 (6.5%) | 1'769 (13.1%) | 2'381 (17.7%) | 2'117 (15.7%) | 1'480 (11.0%) | 1'211 (9.0%) | 1'106 (8.2%) |
| **LV** |  |  |  |  |  |  |  |  |  |  |
| EF | 64 (56, 71) | 64 (56, 71) | 64 (57, 72) | 64 (57, 72) | 65 (57, 72) | 65 (57, 73) | 65 (57, 73) | 65 (57, 73) | 65 (56, 73) | 65 (56, 74) |
| EDV | 136 (100, 174) | 136 (101, 175) | 135 (101, 174) | 134 (99, 173) | 132 (96, 171) | 129 (93, 169) | 126 (90, 165) | 121 (86, 160) | 115 (81, 153) | 109 (75, 146) |
| _bsa | 78 (59, 94) | 77 (60, 94) | 77 (59, 94) | 76 (57, 94) | 74 (55, 93) | 72 (53, 91) | 70 (52, 89) | 68 (50, 87) | 65 (47, 84) | 63 (44, 82) |
| _m<sup>1.7</sup> | 56 (43, 70) | 56 (43, 70) | 56 (43, 70) | 56 (43, 70) | 55 (41, 70) | 54 (40, 69) | 53 (39, 68) | 52 (38, 67) | 50 (36, 65) | 48 (34, 63) |
| _m | 81 (61, 100) | 81 (62, 101) | 81 (62, 102) | 80 (61, 101) | 79 (59, 101) | 78 (57, 99) | 76 (56, 98) | 73 (54, 95) | 71 (51, 92) | 68 (47, 89) |
| ESV | 49 (34, 67) | 49 (34, 67) | 48 (33, 67) | 48 (32, 66) | 47 (31, 65) | 45 (30, 64) | 44 (29, 63) | 42 (27, 61) | 40 (25, 59) | 38 (23, 56) |
| _bsa | 28 (20, 37) | 28 (20, 37) | 27 (19, 37) | 27 (19, 36) | 26 (18, 36) | 25 (17, 35) | 25 (17, 34) | 24 (16, 33) | 23 (15, 32) | 22 (14, 31) |
| _m<sup>1.7</sup> | 20 (14, 27) | 20 (14, 27) | 20 (14, 27) | 20 (14, 27) | 20 (13, 27) | 19 (13, 26) | 19 (12, 26) | 18 (12, 26) | 18 (11, 25) | 17 (11, 25) |
| _m | 29 (21, 40) | 29 (20, 39) | 29 (20, 39) | 28 (20, 39) | 28 (19, 38) | 27 (18, 38) | 27 (18, 37) | 26 (17, 37) | 25 (16, 36) | 24 (15, 34) |
| SV | 86 (63, 112) | 87 (63, 113) | 87 (63, 114) | 86 (62, 113) | 85 (61, 112) | 84 (59, 111) | 82 (57, 109) | 79 (55, 106) | 75 (51, 102) | 71 (48, 97) |
| _bsa | 49 (37, 62) | 49 (37, 62) | 49 (37, 62) | 49 (36, 62) | 48 (35, 61) | 47 (34, 61) | 46 (33, 60) | 44 (31, 58) | 42 (30, 56) | 41 (28, 54) |
| _m<sup>1.7</sup> | 36 (27, 46) | 36 (27, 46) | 36 (27, 46) | 36 (27, 46) | 36 (26, 46) | 35 (25, 46) | 34 (25, 45) | 34 (24, 45) | 33 (23, 43) | 32 (22, 42) |
| _m | 51 (38, 66) | 52 (39, 66) | 52 (38, 67) | 52 (38, 67) | 51 (37, 66) | 50 (36, 66) | 49 (35, 65) | 48 (34, 64) | 46 (32, 61) | 44 (30, 59) |
| CO | 5.4 (3.5, 8) | 5.5 (3.6, 8) | 5.5 (3.6, 8) | 5.5 (3.6, 7.9) | 5.4 (3.5, 7.8) | 5.3 (3.4, 7.7) | 5.2 (3.3, 7.5) | 5 (3.2, 7.3) | 4.8 (3, 7.1) | 4.5 (2.8, 6.8) |
| _bsa | 3.2 (2.1, 4.5) | 3.1 (2.1, 4.4) | 3.1 (2.1, 4.4) | 3.1 (2.1, 4.4) | 3 (2, 4.3) | 3 (1.9, 4.2) | 2.9 (1.9, 4.2) | 2.8 (1.8, 4.1) | 2.7 (1.7, 4) | 2.6 (1.6, 3.8) |
| _m<sup>1.7</sup> | 2.3 (1.5, 3.3) | 2.3 (1.5, 3.3) | 2.3 (1.5, 3.3) | 2.3 (1.5, 3.3) | 2.3 (1.5, 3.2) | 2.2 (1.5, 3.2) | 2.2 (1.4, 3.2) | 2.1 (1.4, 3.1) | 2.1 (1.3, 3) | 2 (1.2, 3) |
| _m | 3.3 (2.1, 4.7) | 3.3 (2.2, 4.7) | 3.3 (2.2, 4.7) | 3.3 (2.2, 4.7) | 3.2 (2.1, 4.6) | 3.2 (2.1, 4.6) | 3.1 (2, 4.5) | 3 (1.9, 4.4) | 2.9 (1.8, 4.3) | 2.8 (1.7, 4.1) |
| Mass | 89 (70, 112) | 91 (70, 116) | 90 (70, 118) | 91 (70, 118) | 91 (69, 117) | 92 (70, 118) | 92 (71, 119) | 91 (70, 118) | 90 (68, 116) | 88 (66, 113) |
| _bsa | 51 (43, 60) | 51 (43, 61) | 51 (42, 61) | 51 (42, 62) | 51 (42, 62) | 51 (42, 62) | 51 (42, 63) | 51 (41, 63) | 51 (41, 63) | 50 (41, 62) |
| _m<sup>1.7</sup> | 37 (30, 45) | 38 (30, 47) | 38 (30, 48) | 38 (30, 48) | 38 (30, 48) | 39 (30, 49) | 39 (30, 50) | 39 (30, 50) | 39 (30, 50) | 39 (30, 50) |
| _m | 53 (43, 65) | 54 (43, 68) | 54 (43, 69) | 54 (43, 69) | 55 (43, 69) | 55 (43, 70) | 55 (43, 71) | 55 (43, 71) | 55 (43, 71) | 55 (42, 69) |
| Wall thickness | 6 (5.3, 6.8) | 6 (5.2, 6.9) | 6 (5.3, 7.1) | 6.1 (5.3, 7.2) | 6.2 (5.3, 7.4) | 6.3 (5.4, 7.5) | 6.4 (5.5, 7.7) | 6.6 (5.5, 7.8) | 6.7 (5.6, 8) | 6.8 (5.7, 8.2) |
| _bsa | 3.4 (3, 3.9) | 3.4 (3, 3.9) | 3.4 (3, 4) | 3.5 (3, 4) | 3.5 (3, 4.1) | 3.5 (3, 4.2) | 3.6 (3.1, 4.3) | 3.7 (3.1, 4.4) | 3.8 (3.2, 4.5) | 3.9 (3.3, 4.7) |
| _m<sup>1.7</sup> | 2.5 (2.1, 2.9) | 2.5 (2.1, 2.9) | 2.5 (2.2, 3) | 2.6 (2.2, 3.1) | 2.6 (2.2, 3.1) | 2.7 (2.2, 3.2) | 2.7 (2.3, 3.3) | 2.8 (2.3, 3.4) | 2.9 (2.4, 3.6) | 3 (2.5, 3.7) |
| _m | 3.5 (3.1, 4) | 3.6 (3.1, 4.1) | 3.6 (3.1, 4.2) | 3.7 (3.2, 4.3) | 3.7 (3.2, 4.4) | 3.8 (3.2, 4.6) | 3.9 (3.3, 4.7) | 4 (3.4, 4.8) | 4.1 (3.4, 4.9) | 4.2 (3.5, 5.1) |
| midseptal | 6.2 (5.3, 7.2) | 6.2 (5.2, 7.4) | 6.3 (5.2, 7.7) | 6.4 (5.3, 7.9) | 6.5 (5.3, 8.2) | 6.7 (5.5, 8.4) | 7 (5.6, 8.7) | 7.2 (5.8, 9) | 7.4 (5.9, 9.2) | 7.6 (6, 9.5) |
| **RV** |  |  |  |  |  |  |  |  |  |  |
| EF | 55 (45, 66) | 56 (45, 66) | 57 (45, 67) | 57 (46, 67) | 57 (46, 68) | 58 (46, 68) | 58 (47, 69) | 58 (47, 69) | 59 (48, 70) | 59 (49, 70) |
| EDV | 151 (108, 198) | 150 (108, 196) | 149 (107, 194) | 147 (105, 192) | 145 (103, 189) | 142 (100, 185) | 137 (97, 181) | 131 (93, 175) | 125 (87, 167) | 117 (81, 158) |
| _bsa | 86 (64, 108) | 85 (64, 107) | 84 (64, 106) | 83 (62, 104) | 81 (60, 103) | 79 (58, 101) | 77 (56, 99) | 74 (54, 96) | 71 (51, 93) | 67 (47, 90) |
| _m<sup>1.7</sup> | 62 (46, 80) | 62 (46, 79) | 62 (46, 79) | 62 (45, 78) | 61 (45, 77) | 59 (44, 76) | 58 (42, 75) | 56 (41, 73) | 54 (39, 71) | 52 (37, 68) |
| _m | 90 (66, 115) | 90 (66, 114) | 89 (65, 113) | 88 (64, 112) | 87 (63, 111) | 85 (62, 109) | 83 (60, 107) | 80 (58, 104) | 76 (55, 100) | 73 (51, 96) |
| ESV | 67 (42, 94) | 66 (41, 93) | 65 (41, 92) | 64 (40, 91) | 62 (39, 89) | 60 (37, 86) | 58 (36, 83) | 55 (34, 80) | 51 (32, 75) | 48 (30, 69) |
| _bsa | 38 (25, 52) | 37 (25, 51) | 37 (24, 50) | 36 (23, 50) | 35 (23, 49) | 33 (22, 47) | 32 (21, 46) | 31 (20, 44) | 29 (19, 41) | 27 (18, 39) |
| _m<sup>1.7</sup> | 28 (18, 38) | 27 (18, 38) | 27 (17, 37) | 27 (17, 37) | 26 (17, 36) | 25 (16, 35) | 24 (16, 34) | 23 (15, 33) | 22 (14, 32) | 21 (14, 30) |
| _m | 40 (25, 55) | 39 (25, 55) | 39 (25, 54) | 38 (24, 53) | 37 (24, 52) | 36 (23, 51) | 35 (22, 49) | 33 (21, 47) | 31 (20, 45) | 29 (19, 42) |
| SV | 83 (58, 111) | 84 (58, 111) | 84 (58, 111) | 83 (57, 111) | 83 (56, 111) | 82 (55, 110) | 80 (54, 108) | 77 (52, 106) | 73 (49, 103) | 70 (46, 98) |
| _bsa | 48 (34, 61) | 48 (34, 62) | 48 (33, 62) | 47 (33, 62) | 46 (32, 62) | 46 (32, 61) | 45 (31, 60) | 43 (30, 59) | 42 (28, 57) | 40 (27, 56) |
| _m<sup>1.7</sup> | 34 (25, 45) | 35 (25, 46) | 35 (25, 46) | 35 (25, 46) | 35 (24, 46) | 34 (24, 45) | 34 (23, 45) | 33 (23, 44) | 32 (22, 44) | 31 (21, 43) |
| _m | 49 (35, 65) | 50 (35, 65) | 50 (35, 66) | 50 (35, 66) | 50 (35, 66) | 49 (34, 65) | 48 (33, 64) | 47 (32, 63) | 45 (31, 62) | 43 (29, 60) |
| CO | 5.3 (3.4, 7.7) | 5.4 (3.4, 7.7) | 5.3 (3.3, 7.8) | 5.3 (3.3, 7.8) | 5.2 (3.3, 7.7) | 5.2 (3.2, 7.7) | 5.1 (3.1, 7.5) | 4.8 (3, 7.3) | 4.7 (2.8, 7) | 4.4 (2.6, 6.7) |
| _bsa | 3 (2, 4.4) | 3 (2, 4.4) | 3 (1.9, 4.4) | 3 (1.9, 4.3) | 3 (1.9, 4.3) | 2.9 (1.8, 4.2) | 2.8 (1.8, 4.2) | 2.7 (1.7, 4.1) | 2.7 (1.6, 4) | 2.6 (1.5, 3.9) |
| _m<sup>1.7</sup> | 2.2 (1.4, 3.2) | 2.2 (1.4, 3.2) | 2.2 (1.4, 3.2) | 2.2 (1.4, 3.2) | 2.2 (1.4, 3.2) | 2.2 (1.4, 3.2) | 2.1 (1.3, 3.1) | 2.1 (1.3, 3.1) | 2 (1.2, 3.1) | 2 (1.2, 3) |
| _m | 3.1 (2, 4.5) | 3.2 (2, 4.5) | 3.2 (2, 4.6) | 3.2 (2, 4.6) | 3.1 (2, 4.6) | 3.1 (2, 4.6) | 3.1 (1.9, 4.5) | 2.9 (1.9, 4.4) | 2.9 (1.7, 4.3) | 2.7 (1.6, 4.2) |

Values are fitted mean (5th quantile, 95th quantile) from an additive model with CMR metric as an outcome and a non-linear smoothing function of continuous age, given by a restricted cubic spline basis and up to 20 knots, averaged by 5-year age group. CMR metrics were indexed to body surface area according to DuBois formula (\_bsa), body height in meters to the power of 1.7 (\_m<sup>1.7</sup>), and body height in meters (\_m). LV: left ventricle; RV: right ventricle; EF: ejection fraction; EDV: end-diastolic volume; ESV: end-systolic volume; SV: stroke volume; CO: cardiac output; BSA: body surface area.

**Table 2b:**
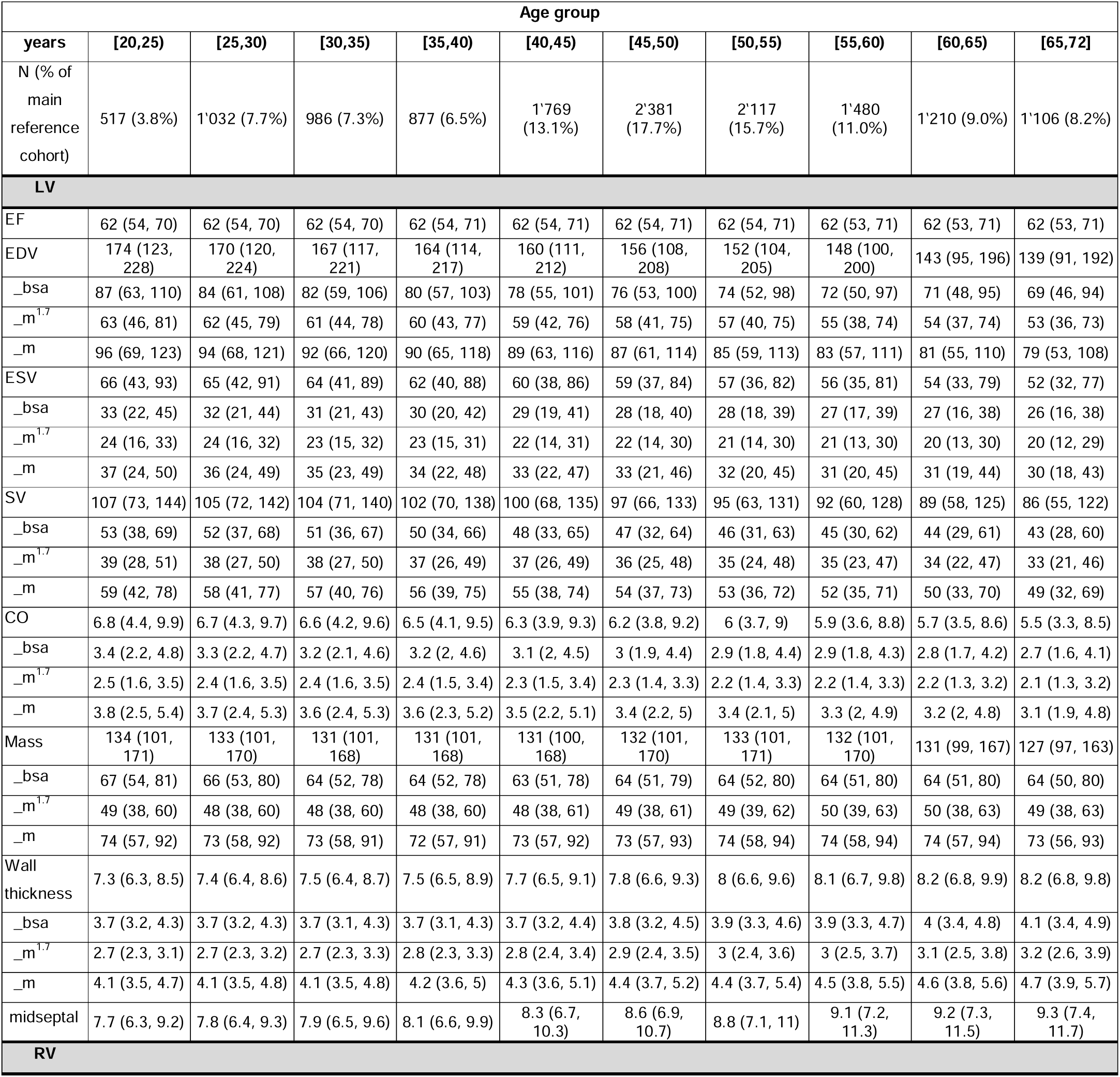
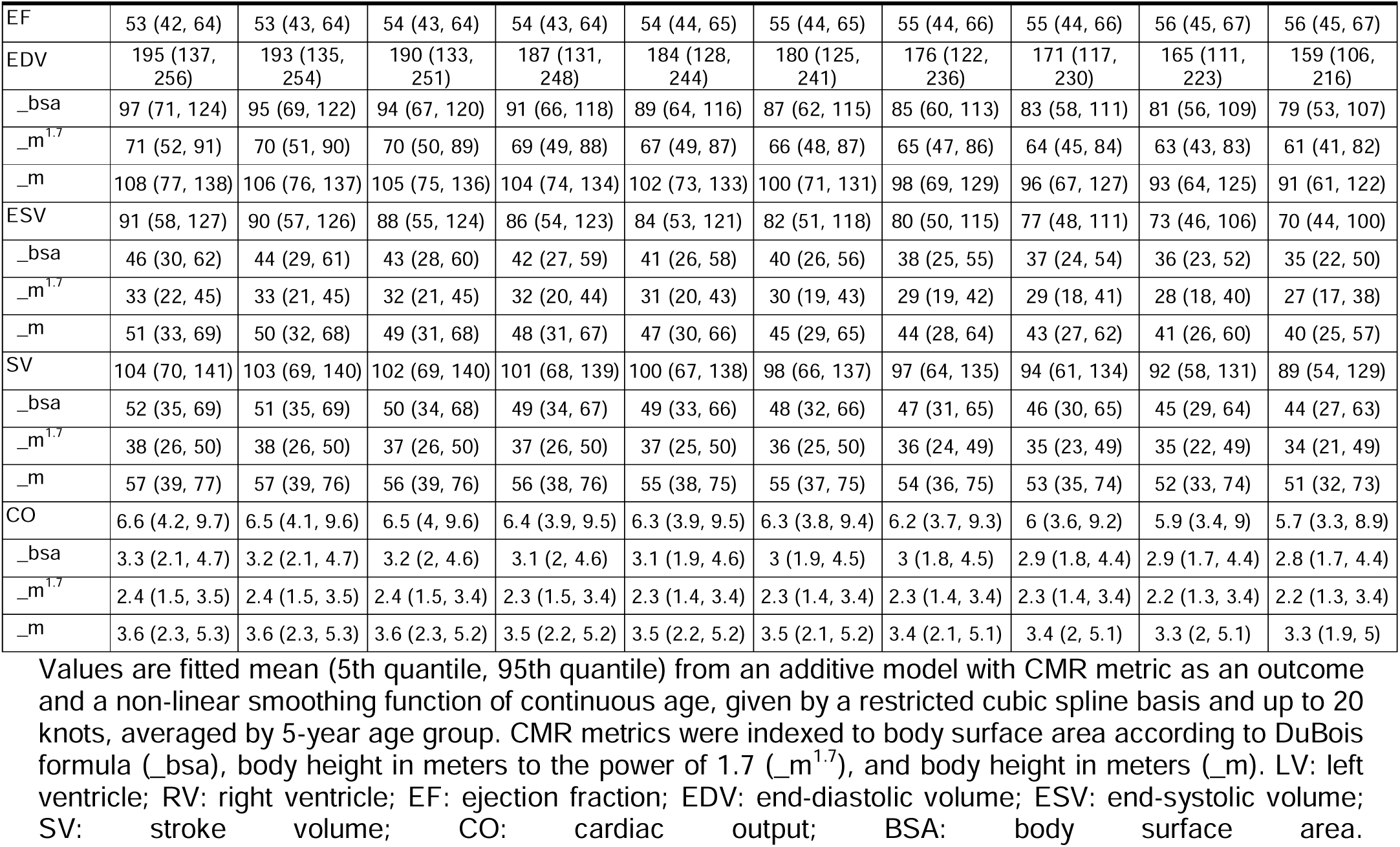
CMR reference ranges for men.

We observed EF preservation across the age range in both sexes while women exhibited higher LVEF (fitted mean 64-65% [Reference range: 56-74%]) and RVEF (55-59% [45–70%]) compared to men (LVEF: 62% [53-71%]; RVEF: 53-56% [42-67%]) across all age groups (**Figure 2**, **Table 2a and 2b**). Men demonstrated larger volumes compared to women, including +30-40% for LVEDV (e.g., 174 vs. 136 ml at 20-25yr) or +30% for RVEDV (195 vs. 151 ml at 20-25yr). After BSA-indexing, differences persisted but were attenuated (e.g., LVEDV_bsa: 87 vs. 78 ml/m² at 20-25yr in men vs women).

Reference ranges for chamber values were shifted to lower values at higher ages for both sexes but the difference was larger for men in absolute terms (e.g., LVEDV difference 35 ml in men vs. 27 ml in women). Myocardial mass showed stable to slightly lower levels at higher ages. Reference ranges for wall thickness were shifted to higher values at higher ages (**Table 2a and 2b**).

### Difference of age effects between women and men

For all CMR metrics, age-outcome associations were significantly different between women and men (all p<0.05, **Supplementary Figure 2**). The effect of age on RVEF and LVEF differed by sex, with a stronger age-related effect in men before approximately 45 years and in women thereafter. For LV volumes, there was an inverted U-shaped sex difference with age effects, with women showing a stronger association with age than men between 35 and 50 years (**Supplementary Figure 2**). RV volumes showed overall similar patterns, but change-pattern started approximately five years earlier.

### Comparison of reference ranges to the Healthy Subcohort

References ranges derived in the healthy subcohort were similar to those in the main reference cohort (**Figure 3**). Although absolute values differed, (e.g. reference ranges for myocardial mass were shifted to lower values, and ranges for LVSV were shifted to higher values in the healthy subcohort), these differences were not clinically relevant, and all ranges overlapped substantially (**Supplementary Figure 1, Supplementary Table 3**). Main differences between main reference cohort and healthy subcohort were present in the older age groups, in particular for mid-septal LV wall thickness (e.g. 9.3 [7.4, 11.7] mm vs. 8.3 [6.9, 10.1] mm for men aged >65 years, **Supplementary Table 3b**) and RV metrics (e.g. 117 [81, 158] ml vs 124 [90, 163] ml RVEDV for women aged >65 years; **Supplementary Table 3a**), in the main reference cohort and healthy subcohort, respectively. These differences can be explained by the selection criteria, since the older age group was disproportionately affected by the exclusion of individuals with CVD risk factors, due to the higher prevalence of these risk factors at higher age.

To further substantiate the role of CVD risk factors for the reference ranges, we compared adjusted R^2^ values from the generalized additive model including smooth terms of age to a model additionally adjusted for the CVD risk factors on the main reference cohort. The largest differences in R^2^, interpretable as the largest contributions of the risk factors to variance in the CMR metric, were observed for mid-septal LV wall thickness (40.6% vs 18.3% in women and 32.9% vs 16.5% in men for the risk factor adjusted model vs the age-only model, respectively). However, R^2^ differences were minor for LVEF (1.1% vs 0.6% in women and 0.6% vs 0.1% in men), LVEDV (14.5% vs 11.7% and 10.1% vs 8.8%), LVESV (11.3% vs 9.0% and 7.4% vs 6.5%), and RVEF (3.1% vs 2.3% and 1.7% vs 1.3%). This small contribution of the risk factors to R^2^ corroborates the similarity of the reference ranges whether individuals with these risk factors are included or not.

## Discussion

In this large, population-based cohort in Germany, we derived age- and sex-specific reference ranges for volumes, mass and function of the left and right ventricle measured by CMR in 24,371 adults without CVD, and in a healthy subcohort of 5,550 individuals without CVD or CVD risk factors. Using generalized additive quantile regression, we provide smooth percentile curves and tabulated 5^th^ to 95^th^ percentile ranges across the adult age span - starting at the age of 20 years. We observed a consistent pattern of lower ventricular volumes with higher age, preservation of ejection fraction across the age range, and marked sex differences in cardiac size and output that were most pronounced at younger ages and attenuated in later life. Taken together, these findings extend and refine existing CMR reference standards and offer population-based reference data acquired on uniform 3T scanner systems within a single national population-based study.

Our results are broadly concordant with prior CMR reference compilations but address several important gaps. The EACVI CMR expert consensus and, more recently, the Society for Cardiovascular Magnetic Resonance reference values paper provided much-needed structure for grading chamber size and function, but they still relied largely on pooled estimates rather than a single, prospective, population-based sample [14, 15]. In contrast, our reference ranges were derived from a uniformly imaged, centrally processed national population-based study with standardized acquisition, automated segmentation and systematic quality control, which likely contributes to the comparatively narrower reference ranges observed for many CMR metrics.

Our data are complementary to the recently published Healthy Hearts Consortium reference ranges, and highlight how population composition and study design influence healthy CMR phenotypes. The Healthy Hearts Consortium assembled CMR scans from multiple cohorts to derive age-, sex- and ethnicity-specific reference ranges and severity grading criteria for ventricular and atrial metrics in carefully phenotyped healthy adults [3]. In NAKO, participants were recruited from the general population, and our main reference cohort only excluded individuals with manifest CVD, while the healthy subcohort additionally excluded those with traditional cardiovascular risk factors. This two-tier approach reflects clinical reality, where CMR is often performed in individuals with risk factors but without established disease. Notably, in our analysis traditional cardiovascular risk factors added little to explain the variance of most CMR metrics.

The age-related patterns observed cross-sectionally in NAKO align with prior work describing structural remodeling of the aging heart. Ventricular volumes decreased with age in both sexes, more steeply in men, while myocardial mass showed a less pronounced association and the EF remained largely preserved. This pattern mirrors the healthy aging phenotype described in previous CMR studies and in conceptual reviews of ventricular remodeling, where healthy aging is characterized by concentric remodeling with relatively stable EF [16].

A key novel aspect of our study is the demonstration of strong age-sex interactions across all major CMR metrics, with the largest sex differences observed at younger ages and a convergence of phenotypes in later life. One plausible explanation for this convergence is the menopausal transition in women. Prior echocardiographic and CMR studies have shown that menopause and earlier age at menopause are associated with smaller LV end-diastolic and stroke volumes and a more concentric LV remodeling pattern, consistent with an acceleration of age-related structural changes in the female heart [17]. In parallel, cardiometabolic risk factors in women tend to catch up with those in men after midlife, which may further contribute to the attenuation of sex differences in cardiac structure [18]. Although we did not explicitly model menopausal status in the present analyses, the observed age-sex interactions are compatible with a scenario in which loss of ovarian hormone exposure and convergence of risk factor profiles in later life reduce the early-life female advantage in cardiac size and function.

Our data also fit well into the emerging framework of biological heart age. Radiomics-based models from UKB have shown that CMR-derived heart age closely tracks chronological age in apparently healthy individuals, with the discrepancy (“heart age gap”) capturing accelerated or decelerated cardiac aging and associating with risk factor burden and outcomes [19]. More recent work has demonstrated differential aging patterns across cardiac chambers and linked larger heart-age gaps to adverse body composition, arterial stiffness and psychosocial factors [20, 21].

From a clinical standpoint, the present NAKO reference ranges may be integrated into routine CMR reporting to contextualize an individual’s measurements within age- and sex-appropriate percentiles. In contrast to static cut-offs, percentile-based interpretation can help distinguish borderline findings in older individuals (e.g. modestly reduced volumes with preserved EF) from clearly pathological values in younger adults. The availability of both a main reference cohort and a strictly healthy subcohort further allows clinicians to decide whether they wish to benchmark their patient against a “real-world” reference (no manifest CVD) or against an idealized, risk-factor-free standard. Finally, these data provide a foundation for NAKO-specific prognostic modeling, including future work on CMR-based heart age and remodeling scores calibrated to hard cardiovascular outcomes.

Our study has several strengths. First, NAKO is a large, national, population-based cohort with standardized recruitment across multiple study centers, enabling derivation of reference ranges that will be applicable to the general adult population in Germany. Second, CMR acquisition was performed on harmonized 3T platforms with uniform protocols, and image analysis relied on an automated segmentation pipeline combined with rigorous quality control, as recently detailed elsewhere, minimizing inter-observer variability and technical heterogeneity. Third, the use of quantile regression allowed us to flexibly model non-linear age relationships and heteroscedasticity and to derive smooth percentile curves across the entire adult age spectrum. This is particularly important because NAKO includes a substantial proportion of younger participants, unlike previous cohorts that focused on middle-aged participants.

Some limitations should be acknowledged. First, despite its size and population-based design, NAKO participants are predominantly of European descent, and we did not perform ethnicity-stratified analyses. Extrapolation of our reference ranges to other ethnic groups should therefore be done with caution and ideally complemented by local data or by multi-ethnic reference datasets such as those emerging from international consortia. Second, our data are cross-sectional; although the observed age patterns are biologically plausible and consistent with prior literature, longitudinal follow-up will be required to confirm individual trajectories of structural cardiac aging and to link deviations from the reference ranges to incident cardiovascular events. Third, we focused on conventional volumetric and functional metrics; future work in NAKO could extend the present reference framework to strain, atrial size and parametric mapping, in line with recent comprehensive CMR reference studies.

In conclusion, using CMR in a large, population-based cohort, we provide age- and sex-specific reference ranges for volumes, mass and function for the left and right ventricle, both in a broad reference population and in a rigorously defined healthy subset. Our data complement and extend existing CMR reference value resources by adding a population-based sample with certain age groups – particularly young adults. These reference ranges may support more nuanced interpretation of CMR studies in clinical practice and serve as a benchmark for future research on structural cardiac aging, biological heart age and ventricular remodeling.

## Data Availability

All data produced in the present study are available upon reasonable request to the authors

## Declarations

### CRediT author statement

Conceptualization: CLS, SR, JSM; Investigation: CLS, CS, PMF, RTS, MH, MR, MFR, JG, CA, KHMH, JSM; Methodology: CLS, SR; Formal analysis: SR; Resources: CLS, CS, PMF, RTS, MH, MR, MFR, MF, JG, CA, VG, KHG, TG, AK, TK, TKü, WL, LJM, KM, AP, TP, HT, HV, KHMH, FB, SR, JSM; Data Curation: CS, RTS, SR; Visualization: SR; Writing—original draft: CLS, CS, SR, JSM; Writing-review and editing: CLS, CS, PMF, RTS, MH, MR, MFR, MF, JG, CA, VG, KHG, TG, AK, TK, TKü, WL, LJM, KM, AP, TP, HT, HV, KHMH, FB, SR, JSM

## Acknowledgments

This project was conducted with data (application no. NAKO-249, NAKO-744) from the German National Cohort (NAKO). We thank all participants who took part in the NAKO study and the staff of this research initiative.

## Supplementary Material to

**Age- and Sex-specific Reference Ranges for Cardiac Function and Structure in Germany: Cardiovascular Magnetic Resonance Imaging (CMR) in the German National Cohort (NAKO)**

### Supplementary Tables

**Supplementary Table 1:** Participant characteristics according to MR imaging site.

|  | Augsburg | Mannheim | Essen | Berlin | Neubrandenburg |
| --- | --- | --- | --- | --- | --- |
|  | N = 5'007 | N = 4'746 | N = 4'708 | N = 4'722 | N = 5'188 |
| Age, years | 48.1 ± 12.2 | 47.0 ± 12.6 | 46.7 ± 12.5 | 46.5 ± 12.0 | 49.4 ± 11.4 |
| Women | 2'229 (44.5%) | 2'063 (43.5%) | 2'065 (43.9%) | 2'034 (43.1%) | 2'505 (48.3%) |
| Height, cm | 172.1 ± 9.3 | 172.9 ± 9.7 | 173.9 ± 9.6 | 174.1 ± 9.5 | 172.3 ± 9.5 |
| Weight, kg | 78.9 ± 16.1 | 78.5 ± 16.0 | 80.1 ± 16.4 | 77.7 ± 15.1 | 81.1 ± 16.4 |
| BMI, kg/m<sup>2</sup> | 26.6 ± 4.7 | 26.2 ± 4.6 | 26.4 ± 4.7 | 25.6 ± 4.3 | 27.2 ± 4.7 |
| BSA, m<sup>2</sup> | 1.91 ± 0.22 | 1.92 ± 0.22 | 1.94 ± 0.22 | 1.92 ± 0.21 | 1.94 ± 0.22 |
| SCORE2, % | 2.6 [1.1, 5.2] | 2.4 [1.0, 4.8] | 2.5 [1.0, 5.1] | 2.5 [1.1, 4.9] | 3.1 [1.4, 5.8] |
| FRS10, % | 7.9 [3.3, 16.2] | 7.2 [3.0, 15.1] | 7.5 [3.0, 15.7] | 7.6 [3.2, 15.3] | 9.8 [4.5, 18.6] |
| FRS30, % | 26.9 [13.5, 43.2] | 25.2 [12.5, 41.4] | 25.6 [12.4, 43.3] | 26.4 [13.4, 43.1] | 32.1 [17.6, 49.1] |
| Hypertension | 1'872 (37.4%) | 1'507 (31.8%) | 1'656 (35.2%) | 1'505 (31.9%) | 2'393 (46.1%) |
| Diabetes | 215 (4.3%) | 188 (4.0%) | 227 (4.8%) | 159 (3.4%) | 292 (5.6%) |
| Obesity | 1'011 (20.2%) | 836 (17.6%) | 918 (19.5%) | 648 (13.7%) | 1'244 (24.0%) |
| Hypercholesterolemia | 1'704 (34.0%) | 1'371 (28.9%) | 1'391 (29.5%) | 1'537 (32.5%) | 1'742 (33.6%) |
| Ever Smoking | 2'399 (47.9%) | 2'155 (45.4%) | 2'245 (47.7%) | 2'278 (48.2%) | 2'662 (51.3%) |
| Inclusion in healthy subcohort | 1'085 (21.7 %) | 1'293 (27.2 %) | 1'182 (25.1%) | 1'140 (24.1%) | 850 (16.4%) |

Values are presented as mean ± standard deviation, or median [Q1, Q3] for continuous variables, and counts with percentages for categorical variables. BMI: Body mass index; BSA: body surface area; FRS: Framingham Risk Score.

**Supplementary Table 2:** CMR metrics in the main reference cohort and healthy subcohort.

|  | Main reference cohort | Healthy subcohort | Main reference cohort, Women | Healthy subcohort, Women | Main reference cohort, Men | Healthy subcohort, Men |
| --- | --- | --- | --- | --- | --- | --- |
| N | N = 24'371 | N = 5'550 | N = 10'896 (44.7%) | N = 2'802 (50.5%) | N = 13'475 (55.3%) | N = 2'748 (49.5%) |
| **LV** |  |  |  |  |  |  |
| EF, % | 63.4 ± 5.3 | 63.6 ± 5.0 | 64.8 ± 4.9 | 64.8 ± 4.8 | 62.3 ± 5.3 | 62.4 ± 5.0 |
| EDV, ml | 142.3 ± 32.5 | 146.1 ± 32.7 | 125.9 ± 24.0 | 128.6 ± 22.4 | 155.6 ± 32.4 | 164.0 ± 31.8 |
| _bsa | 73.8 ± 14.0 | 78.5 ± 13.5 | 70.9 ± 12.1 | 74.7 ± 11.2 | 76.1 ± 15.0 | 82.4 ± 14.5 |
| _m<sup>1.7</sup> | 55.7 ± 10.4 | 56.9 ± 9.9 | 53.3 ± 9.0 | 53.8 ± 8.2 | 57.6 ± 11.0 | 60.0 ± 10.5 |
| _m | 81.9 ± 16.3 | 83.8 ± 16.0 | 75.9 ± 13.3 | 77.0 ± 12.2 | 86.7 ± 16.9 | 90.7 ± 16.3 |
| ESV, ml | 52.4 ± 15.1 | 53.4 ± 15.1 | 44.5 ± 10.9 | 45.3 ± 10.2 | 58.8 ± 15.1 | 61.7 ± 14.8 |
| _bsa | 27.1 ± 6.7 | 28.6 ± 6.6 | 25.0 ± 5.6 | 26.3 ± 5.4 | 28.7 ± 7.0 | 31.0 ± 6.9 |
| _m<sup>1.7</sup> | 20.5 ± 5.0 | 20.7 ± 4.8 | 18.8 ± 4.3 | 18.9 ± 3.9 | 21.8 ± 5.2 | 22.6 ± 5.0 |
| _m | 30.1 ± 7.8 | 30.6 ± 7.7 | 26.8 ± 6.2 | 27.1 ± 5.7 | 32.8 ± 8.0 | 34.1 ± 7.8 |
| SV, ml | 90.0 ± 20.8 | 92.7 ± 20.9 | 81.5 ± 16.3 | 83.3 ± 15.5 | 96.8 ± 21.5 | 102.3 ± 21.2 |
| _bsa | 46.7 ± 9.5 | 49.9 ± 9.1 | 45.9 ± 8.5 | 48.4 ± 8.1 | 47.4 ± 10.1 | 51.4 ± 9.9 |
| _m<sup>1.7</sup> | 35.3 ± 7.0 | 36.1 ± 6.7 | 34.5 ± 6.3 | 34.8 ± 5.9 | 35.9 ± 7.4 | 37.5 ± 7.2 |
| _m | 51.8 ± 10.7 | 53.2 ± 10.5 | 49.1 ± 9.2 | 49.8 ± 8.7 | 54.0 ± 11.3 | 56.6 ± 11.1 |
| CO, l/min | 5.7 ± 1.7 | 5.9 ± 1.6 | 5.2 ± 1.4 | 5.3 ± 1.3 | 6.2 ± 1.7 | 6.4 ± 1.7 |
| _bsa | 3.0 ± 0.8 | 3.1 ± 0.8 | 2.9 ± 0.8 | 3.1 ± 0.7 | 3.0 ± 0.8 | 3.2 ± 0.8 |
| _m<sup>1.7</sup> | 2.2 ± 0.6 | 2.3 ± 0.6 | 2.2 ± 0.6 | 2.2 ± 0.5 | 2.3 ± 0.6 | 2.4 ± 0.6 |
| _m | 3.3 ± 0.9 | 3.4 ± 0.9 | 3.1 ± 0.8 | 3.2 ± 0.8 | 3.4 ± 0.9 | 3.6 ± 0.9 |
| Mass, g | 113.4 ± 27.5 | 106.4 ± 26.1 | 90.9 ± 14.8 | 86.4 ± 12.6 | 131.6 ± 21.2 | 126.9 ± 19.7 |
| _bsa | 58.4 ± 10.1 | 56.9 ± 9.8 | 51.1 ± 6.3 | 50.2 ± 5.8 | 64.2 ± 8.6 | 63.7 ± 8.3 |
| _m<sup>1.7</sup> | 44.2 ± 8.4 | 41.3 ± 7.6 | 38.6 ± 5.9 | 36.2 ± 4.6 | 48.8 ± 7.3 | 46.5 ± 6.5 |
| _m | 65.1 ± 13.6 | 60.9 ± 12.6 | 54.9 ± 8.4 | 51.7 ± 6.8 | 73.4 ± 11.1 | 70.3 ± 10.0 |
| Wall thickness, mm | 7.2 ± 1.1 | 6.7 ± 0.9 | 6.4 ± 0.7 | 6.0 ± 0.5 | 7.8 ± 0.9 | 7.3 ± 0.7 |
| _bsa | 3.7 ± 0.4 | 3.6 ± 0.3 | 3.6 ± 0.4 | 3.5 ± 0.3 | 3.8 ± 0.4 | 3.7 ± 0.4 |
| _m<sup>1.7</sup> | 2.8 ± 0.4 | 2.6 ± 0.3 | 2.7 ± 0.4 | 2.5 ± 0.2 | 2.9 ± 0.4 | 2.7 ± 0.3 |
| _m | 4.1 ± 0.6 | 3.8 ± 0.4 | 3.9 ± 0.5 | 3.6 ± 0.3 | 4.4 ± 0.5 | 4.1 ± 0.4 |
| midseptal | 7.8 ± 1.4 | 7.0 ± 1.1 | 6.9 ± 1.0 | 6.3 ± 0.7 | 8.6 ± 1.3 | 7.8 ± 0.9 |
| **RV** |  |  |  |  |  |  |
| EF, % | 56.1 ± 6.9 | 55.7 ± 6.9 | 57.8 ± 6.7 | 57.2 ± 6.7 | 54.6 ± 6.7 | 54.1 ± 6.6 |
| EDV, ml | 160.3 ± 38.5 | 164.9 ± 39.2 | 137.6 ± 27.4 | 142.4 ± 26.5 | 178.7 ± 36.2 | 188.0 ± 36.5 |
| _bsa | 83.0 ± 16.3 | 88.5 ± 16.2 | 77.5 ± 14.1 | 82.7 ± 13.4 | 87.4 ± 16.7 | 94.4 ± 16.6 |
| _m<sup>1.7</sup> | 62.7 ± 12.1 | 64.1 ± 12.0 | 58.3 ± 10.3 | 59.5 ± 9.8 | 66.2 ± 12.2 | 68.8 ± 12.2 |
| _m | 92.2 ± 19.2 | 94.5 ± 19.3 | 82.9 ± 15.2 | 85.2 ± 14.6 | 99.6 ± 18.9 | 104.0 ± 18.8 |
| ESV, ml | 70.9 ± 22.0 | 73.6 ± 22.5 | 58.2 ± 15.5 | 61.2 ± 15.9 | 81.2 ± 21.0 | 86.3 ± 21.2 |
| _bsa | 36.6 ± 9.7 | 39.3 ± 10.0 | 32.8 ± 8.2 | 35.5 ± 8.4 | 39.7 ± 9.8 | 43.3 ± 9.9 |
| _m<sup>1.7</sup> | 27.6 ± 7.3 | 28.5 ± 7.3 | 24.6 ± 6.0 | 25.5 ± 6.1 | 30.1 ± 7.3 | 31.6 ± 7.3 |
| _m | 40.7 ± 11.4 | 42.1 ± 11.6 | 35.1 ± 8.8 | 36.6 ± 9.0 | 45.2 ± 11.2 | 47.7 ± 11.2 |
| SV, ml | 89.4 ± 22.4 | 91.4 ± 22.5 | 79.4 ± 17.5 | 81.2 ± 16.6 | 97.5 ± 22.5 | 101.7 ± 23.0 |
| _bsa | 46.4 ± 10.2 | 49.2 ± 10.2 | 44.8 ± 9.4 | 47.2 ± 9.0 | 47.7 ± 10.7 | 51.1 ± 10.9 |
| _m<sup>1.7</sup> | 35.0 ± 7.6 | 35.6 ± 7.4 | 33.6 ± 6.9 | 34.0 ± 6.5 | 36.2 ± 7.9 | 37.2 ± 8.0 |
| _m | 51.5 ± 11.6 | 52.4 ± 11.5 | 47.9 ± 10.0 | 48.6 ± 9.4 | 54.4 ± 12.0 | 56.3 ± 12.2 |
| CO, l/min | 5.7 ± 1.7 | 5.8 ± 1.7 | 5.0 ± 1.4 | 5.1 ± 1.3 | 6.2 ± 1.8 | 6.4 ± 1.8 |
| _bsa | 3.0 ± 0.8 | 3.1 ± 0.8 | 2.8 ± 0.8 | 3.0 ± 0.7 | 3.0 ± 0.9 | 3.2 ± 0.9 |
| _m<sup>1.7</sup> | 2.2 ± 0.6 | 2.2 ± 0.6 | 2.1 ± 0.6 | 2.1 ± 0.5 | 2.3 ± 0.6 | 2.3 ± 0.6 |
| _m | 3.3 ± 0.9 | 3.3 ± 0.9 | 3.0 ± 0.8 | 3.1 ± 0.8 | 3.5 ± 1.0 | 3.5 ± 1.0 |

Values are mean ± standard deviation. CMR metrics were indexed to body surface area according to DuBois formula (\_bsa), body height in meters to the power of 1.7 (\_m<sup>1.7</sup>), and body height in meters (\_m). Abbreviations: LV: left ventricle; RV: right ventricle; EF: ejection fraction; ESV: end-systolic volume; EDV: end-diastolic volume; SV: stroke volume; CO: cardiac output.

**Supplementary Table 3a:** CMR reference ranges for women, based on the healthy subcohort.

| Age group |  |  |  |  |  |  |  |  |  |  |
| --- | --- | --- | --- | --- | --- | --- | --- | --- | --- | --- |
| years | [20,25) | [25,30) | [30,35) | [35,40) | [40,45) | [45,50) | [50,55) | [55,60) | [60,65) | [65,72] |
| N (% of healthy subcohort) | 248 (8.9%) | 380 (13.6%) | 260 (9.3%) | 181 (6.5%) | 420 (15.0%) | 545 (19.5%) | 379 (13.5%) | 181 (6.5%) | 141 (5.0%) | 67 (2.4%) |
| **LV** |  |  |  |  |  |  |  |  |  |  |
| EF | 64 (56, 71) | 64 (57, 71) | 65 (57, 72) | 65 (57, 72) | 65 (57, 73) | 65 (57, 73) | 65 (57, 73) | 65 (56, 74) | 65 (56, 74) | 65 (55, 74) |
| EDV | 134 (99, 169) | 133 (98, 171) | 133 (98, 172) | 132 (96, 171) | 131 (94, 170) | 128 (92, 166) | 125 (90, 161) | 121 (87, 155) | 116 (84, 149) | 112 (81, 144) |
| _bsa | 78 (61, 94) | 78 (60, 95) | 77 (59, 95) | 77 (58, 95) | 76 (57, 94) | 74 (55, 92) | 73 (54, 91) | 71 (52, 89) | 69 (51, 87) | 67 (49, 85) |
| _m<sup>1.7</sup> | 55 (43, 67) | 55 (43, 68) | 55 (42, 69) | 55 (41, 69) | 54 (41, 69) | 54 (40, 68) | 53 (39, 66) | 51 (39, 65) | 50 (38, 63) | 49 (37, 62) |
| _m | 79 (61, 97) | 79 (60, 99) | 79 (60, 100) | 79 (59, 100) | 78 (58, 99) | 77 (57, 97) | 75 (56, 95) | 73 (54, 92) | 71 (53, 90) | 69 (51, 87) |
| ESV | 48 (34, 64) | 48 (33, 64) | 47 (32, 65) | 47 (31, 65) | 46 (30, 65) | 45 (29, 64) | 44 (28, 62) | 42 (28, 59) | 41 (27, 57) | 39 (26, 54) |
| _bsa | 28 (21, 37) | 28 (20, 37) | 27 (20, 36) | 27 (19, 36) | 26 (18, 36) | 26 (18, 36) | 25 (17, 35) | 25 (17, 34) | 24 (16, 33) | 23 (15, 32) |
| _m<sup>1.7</sup> | 20 (14, 26) | 20 (14, 26) | 20 (14, 26) | 19 (13, 26) | 19 (13, 26) | 19 (13, 26) | 18 (12, 25) | 18 (12, 25) | 17 (12, 24) | 17 (11, 23) |
| _m | 29 (21, 37) | 28 (20, 37) | 28 (20, 38) | 28 (19, 38) | 27 (19, 38) | 27 (18, 37) | 26 (18, 36) | 25 (17, 35) | 25 (16, 34) | 24 (16, 33) |
| SV | 85 (61, 110) | 86 (62, 112) | 86 (61, 113) | 86 (61, 112) | 85 (60, 111) | 83 (59, 109) | 81 (58, 107) | 79 (56, 104) | 76 (53, 100) | 73 (50, 97) |
| _bsa | 50 (38, 62) | 50 (38, 63) | 50 (37, 63) | 50 (37, 63) | 49 (36, 62) | 48 (35, 62) | 47 (34, 61) | 46 (33, 60) | 45 (32, 59) | 43 (31, 58) |
| _m<sup>1.7</sup> | 35 (26, 45) | 36 (26, 45) | 36 (26, 46) | 36 (26, 46) | 35 (26, 46) | 35 (25, 45) | 34 (25, 44) | 33 (24, 43) | 33 (23, 42) | 32 (22, 41) |
| _m | 51 (38, 64) | 51 (38, 65) | 51 (38, 66) | 51 (37, 66) | 51 (37, 66) | 50 (36, 65) | 49 (35, 64) | 48 (34, 62) | 46 (33, 60) | 45 (31, 58) |
| CO | 5.4 (3.5, 7.9) | 5.5 (3.5, 7.8) | 5.3 (3.5, 7.8) | 5.5 (3.5, 7.8) | 5.3 (3.5, 7.7) | 5.4 (3.4, 7.6) | 5.2 (3.3, 7.4) | 4.9 (3.2, 7.2) | 4.6 (3, 6.9) | 4.6 (2.8, 6.6) |
| _bsa | 3.1 (2.1, 4.5) | 3.2 (2.1, 4.5) | 3.1 (2.1, 4.4) | 3.2 (2.1, 4.4) | 3.1 (2.1, 4.3) | 3.1 (2.1, 4.3) | 3 (2, 4.2) | 2.9 (1.9, 4.2) | 2.8 (1.8, 4.1) | 2.8 (1.7, 4.1) |
| _m<sup>1.7</sup> | 2.2 (1.5, 3.2) | 2.3 (1.5, 3.2) | 2.2 (1.5, 3.2) | 2.3 (1.5, 3.2) | 2.2 (1.5, 3.1) | 2.2 (1.5, 3.1) | 2.2 (1.5, 3.1) | 2.1 (1.4, 3) | 2 (1.3, 3) | 2 (1.3, 2.9) |
| _m | 3.2 (2.1, 4.6) | 3.3 (2.1, 4.6) | 3.2 (2.2, 4.6) | 3.3 (2.1, 4.6) | 3.2 (2.1, 4.5) | 3.2 (2.1, 4.5) | 3.1 (2.1, 4.4) | 3 (2, 4.3) | 2.8 (1.9, 4.2) | 2.8 (1.8, 4) |
| Mass | 87 (70, 111) | 87 (69, 111) | 87 (69, 111) | 87 (68, 110) | 87 (68, 109) | 86 (67, 109) | 86 (66, 108) | 85 (66, 106) | 83 (65, 105) | 82 (65, 103) |
| _bsa | 51 (43, 59) | 51 (43, 60) | 51 (42, 61) | 50 (42, 61) | 50 (41, 61) | 50 (41, 60) | 50 (41, 60) | 50 (40, 60) | 49 (40, 60) | 49 (39, 60) |
| _m<sup>1.7</sup> | 36 (30, 43) | 36 (30, 44) | 36 (30, 45) | 36 (29, 45) | 36 (29, 45) | 36 (29, 45) | 36 (29, 44) | 36 (29, 44) | 36 (29, 44) | 36 (29, 44) |
| _m | 52 (43, 63) | 52 (42, 64) | 52 (42, 64) | 52 (42, 64) | 52 (41, 64) | 52 (41, 64) | 52 (41, 64) | 51 (41, 63) | 51 (40, 62) | 50 (40, 62) |
| Wall | 5.9 (5.2, 6.6) | 5.9 (5.2, 6.7) | 5.9 (5.2, 6.7) | 5.9 (5.2, 6.8) | 6 (5.2, 6.9) | 6 (5.3, 6.9) | 6.1 (5.3, 7) | 6.2 (5.3, 7) | 6.2 (5.3, 7.1) | 6.3 (5.4, 7.2) |

| thickness |  |  |  |  |  |  |  |  |  |  |
| --- | --- | --- | --- | --- | --- | --- | --- | --- | --- | --- |
| _bsa | 3.4 (3, 3.9) | 3.4 (3, 3.9) | 3.4 (3, 3.9) | 3.5 (3, 4) | 3.5 (3, 4) | 3.5 (3, 4.1) | 3.5 (3, 4.1) | 3.6 (3.1, 4.2) | 3.7 (3.2, 4.3) | 3.8 (3.3, 4.4) |
| _m<sup>1.7</sup> | 2.4 (2.1, 2.8) | 2.4 (2.1, 2.8) | 2.5 (2.1, 2.8) | 2.5 (2.1, 2.9) | 2.5 (2.1, 2.9) | 2.5 (2.2, 3) | 2.6 (2.2, 3) | 2.6 (2.2, 3.1) | 2.7 (2.3, 3.2) | 2.8 (2.3, 3.3) |
| _m | 3.5 (3.1, 4) | 3.5 (3.1, 4) | 3.5 (3.1, 4) | 3.5 (3.1, 4) | 3.6 (3.1, 4.1) | 3.6 (3.2, 4.2) | 3.7 (3.2, 4.2) | 3.7 (3.2, 4.3) | 3.8 (3.3, 4.4) | 3.9 (3.3, 4.5) |
| midseptal | 6.1 (5.2, 7) | 6 (5.2, 7.1) | 6.1 (5.1, 7.2) | 6.1 (5.1, 7.2) | 6.2 (5.2, 7.3) | 6.3 (5.3, 7.5) | 6.4 (5.4, 7.7) | 6.6 (5.5, 7.9) | 6.7 (5.6, 8.1) | 6.9 (5.6, 8.3) |
| **RV** |  |  |  |  |  |  |  |  |  |  |
| EF | 56 (45, 66) | 57 (46, 67) | 57 (46, 67) | 57 (46, 68) | 57 (46, 68) | 58 (46, 68) | 58 (46, 68) | 58 (46, 68) | 58 (46, 69) | 59 (46, 69) |
| EDV | 149 (108, 197) | 148 (106, 195) | 148 (104, 193) | 146 (102, 191) | 144 (100, 189) | 141 (98, 186) | 138 (97, 182) | 134 (95, 175) | 129 (92, 169) | 124 (90, 163) |
| _bsa | 87 (65, 109) | 86 (65, 108) | 86 (64, 107) | 85 (63, 107) | 83 (61, 105) | 82 (60, 104) | 80 (59, 102) | 78 (57, 99) | 76 (56, 97) | 74 (54, 95) |
| _m<sup>1.7</sup> | 62 (46, 79) | 62 (45, 78) | 61 (45, 78) | 61 (44, 77) | 60 (44, 76) | 59 (43, 75) | 58 (42, 74) | 57 (42, 72) | 55 (41, 70) | 54 (40, 69) |
| _m | 89 (66, 114) | 88 (65, 113) | 88 (64, 112) | 87 (63, 111) | 86 (62, 110) | 85 (61, 109) | 83 (60, 107) | 81 (59, 103) | 78 (58, 100) | 76 (57, 97) |
| ESV | 66 (40, 94) | 65 (40, 93) | 64 (39, 92) | 63 (38, 91) | 62 (37, 90) | 60 (37, 87) | 58 (36, 84) | 56 (35, 81) | 54 (34, 77) | 51 (33, 73) |
| _bsa | 38 (25, 53) | 38 (24, 52) | 37 (24, 51) | 37 (23, 51) | 36 (23, 50) | 35 (22, 49) | 34 (22, 47) | 33 (21, 46) | 32 (21, 45) | 31 (20, 44) |
| _m<sup>1.7</sup> | 27 (17, 38) | 27 (17, 38) | 27 (17, 37) | 26 (16, 37) | 26 (16, 36) | 25 (16, 35) | 24 (16, 35) | 24 (15, 34) | 23 (15, 33) | 22 (15, 32) |
| _m | 39 (25, 55) | 39 (24, 54) | 38 (24, 54) | 38 (23, 53) | 37 (23, 52) | 36 (22, 51) | 35 (22, 50) | 34 (22, 48) | 33 (21, 46) | 31 (21, 44) |
| SV | 83 (57, 110) | 84 (58, 110) | 84 (58, 109) | 83 (57, 109) | 82 (56, 109) | 82 (56, 109) | 80 (54, 108) | 77 (53, 106) | 74 (51, 104) | 73 (49, 101) |
| _bsa | 48 (35, 62) | 49 (35, 62) | 49 (34, 63) | 48 (34, 62) | 47 (33, 62) | 47 (33, 62) | 47 (32, 61) | 45 (31, 61) | 44 (31, 60) | 44 (30, 59) |
| _m<sup>1.7</sup> | 34 (25, 45) | 35 (25, 45) | 35 (25, 45) | 35 (24, 45) | 34 (24, 45) | 34 (24, 45) | 34 (23, 44) | 33 (23, 44) | 32 (22, 44) | 32 (22, 44) |
| _m | 49 (35, 65) | 50 (35, 65) | 50 (35, 65) | 49 (35, 65) | 49 (34, 64) | 49 (34, 64) | 48 (33, 64) | 47 (33, 63) | 45 (32, 62) | 45 (31, 61) |
| CO | 5.3 (3.3, 7.5) | 5.3 (3.3, 7.5) | 5.2 (3.3, 7.6) | 5.3 (3.3, 7.6) | 5.1 (3.3, 7.6) | 5.2 (3.2, 7.6) | 5.2 (3.2, 7.5) | 4.9 (3.1, 7.2) | 4.5 (2.9, 6.9) | 4.6 (2.7, 6.6) |
| _bsa | 3.1 (2, 4.3) | 3.1 (2, 4.3) | 3.1 (2, 4.4) | 3 (2, 4.3) | 3 (1.9, 4.3) | 3 (1.9, 4.3) | 3 (1.9, 4.2) | 2.8 (1.8, 4.2) | 2.7 (1.7, 4.1) | 2.8 (1.7, 4) |
| _m<sup>1.7</sup> | 2.2 (1.4, 3) | 2.2 (1.4, 3.1) | 2.2 (1.4, 3.1) | 2.2 (1.4, 3.1) | 2.1 (1.4, 3.1) | 2.2 (1.4, 3.2) | 2.2 (1.4, 3.1) | 2.1 (1.3, 2.9) | 2 (1.3, 2.9) | 2 (1.2, 2.9) |
| _m | 3.1 (2, 4.4) | 3.1 (2, 4.5) | 3.1 (2, 4.5) | 3.1 (2, 4.5) | 3.1 (2, 4.4) | 3.1 (2, 4.6) | 3.1 (1.9, 4.5) | 2.9 (1.9, 4.2) | 2.8 (1.8, 4.1) | 2.8 (1.7, 4.1) |

Values are fitted mean (5th quantile, 95th quantile) from an additive model with CMR metric as an outcome and a non-linear smoothing function of continuous age, given by a restricted cubic spline basis and up to 20 knots, averaged by 5-year age group. CMR metrics were indexed to body surface area according to DuBois formula (\_bsa), body height in meters to the power of 1.7 (\_m<sup>1.7</sup>), and body height in meters (\_m). LV: left ventricle; RV: right ventricle; EF: ejection fraction; EDV: end-diastolic volume; ESV: end-systolic volume; SV: stroke volume; CO: cardiac output; BSA: body surface area.

**Supplementary Table 3b:** CMR reference ranges for men, based on the healthy subcohort.

| Age group |  |  |  |  |  |  |  |  |  |  |
| --- | --- | --- | --- | --- | --- | --- | --- | --- | --- | --- |
| years | [20,25) | [25,30) | [30,35) | [35,40) | [40,45) | [45,50) | [50,55) | [55,60) | [60,65) | [65,72] |
| N (% of healthy subcohort) | 281 (10.2%) | 470 (17.1%) | 333 (12.1%) | 227 (8.3%) | 407 (14.8%) | 434 (15.8%) | 313 (11.4%) | 151 (5.5%) | 72 (2.6%) | 60 (2.2%) |
| **LV** |  |  |  |  |  |  |  |  |  |  |
| EF | 62 (54, 70) | 62 (54, 70) | 62 (54, 70) | 62 (54, 71) | 63 (55, 71) | 63 (55, 71) | 63 (54, 71) | 63 (54, 72) | 63 (54, 72) | 63 (54, 72) |
| EDV | 172 (122, 228) | 172 (121, 226) | 170 (119, 223) | 166 (117, 219) | 163 (114, 216) | 160 (111, 213) | 158 (107, 209) | 153 (104, 202) | 146 (99, 195) | 139 (95, 186) |
| _bsa | 88 (65, 111) | 87 (63, 109) | 85 (62, 108) | 83 (60, 106) | 82 (58, 105) | 80 (57, 103) | 78 (55, 102) | 77 (53, 100) | 75 (52, 99) | 73 (50, 97) |
| _m<sup>1.7</sup> | 63 (46, 81) | 63 (45, 80) | 62 (45, 79) | 61 (44, 78) | 59 (43, 77) | 59 (42, 76) | 58 (41, 75) | 57 (40, 74) | 55 (38, 73) | 53 (36, 72) |
| _m | 95 (68, 123) | 95 (68, 122) | 94 (67, 121) | 92 (66, 119) | 90 (64, 117) | 89 (63, 116) | 88 (61, 114) | 85 (59, 111) | 82 (57, 109) | 78 (54, 105) |
| ESV | 67 (43, 92) | 65 (42, 91) | 64 (41, 89) | 63 (40, 88) | 61 (39, 86) | 60 (38, 84) | 58 (37, 82) | 56 (36, 80) | 54 (35, 76) | 52 (34, 73) |
| _bsa | 34 (23, 45) | 33 (22, 44) | 32 (22, 44) | 31 (21, 43) | 31 (20, 42) | 30 (20, 41) | 29 (19, 40) | 28 (18, 40) | 27 (18, 39) | 27 (17, 38) |
| _m<sup>1.7</sup> | 24 (16, 33) | 24 (16, 32) | 23 (16, 32) | 23 (15, 31) | 22 (15, 31) | 22 (14, 30) | 21 (14, 30) | 21 (14, 29) | 20 (13, 28) | 20 (13, 28) |
| _m | 37 (24, 50) | 36 (24, 49) | 35 (23, 48) | 35 (23, 47) | 34 (22, 46) | 33 (22, 46) | 32 (21, 45) | 31 (20, 44) | 31 (20, 43) | 30 (19, 42) |
| SV | 106 (72, 145) | 106 (73, 143) | 105 (73, 141) | 104 (72, 139) | 102 (70, 137) | 101 (68, 135) | 99 (66, 133) | 96 (62, 131) | 92 (59, 129) | 87 (55, 127) |
| _bsa | 54 (38, 70) | 54 (38, 69) | 53 (38, 69) | 52 (37, 68) | 51 (36, 67) | 50 (35, 67) | 49 (33, 66) | 48 (32, 65) | 48 (30, 65) | 47 (29, 64) |
| _m<sup>1.7</sup> | 39 (27, 51) | 39 (27, 51) | 38 (27, 50) | 38 (27, 50) | 37 (26, 49) | 37 (26, 49) | 37 (25, 48) | 36 (24, 48) | 35 (23, 47) | 33 (22, 47) |
| _m | 59 (41, 78) | 59 (41, 77) | 58 (41, 77) | 57 (41, 76) | 56 (40, 75) | 56 (38, 74) | 55 (37, 73) | 54 (36, 72) | 52 (34, 71) | 49 (33, 70) |
| CO | 6.7 (4.4, 9.9) | 6.7 (4.3, 9.8) | 6.6 (4.2, 9.7) | 6.5 (4.1, 9.6) | 6.4 (4, 9.5) | 6.4 (3.9, 9.4) | 6.3 (3.8, 9.2) | 6.1 (3.7, 8.9) | 5.8 (3.6, 8.6) | 5.5 (3.5, 8.2) |
| _bsa | 3.4 (2.3, 4.9) | 3.4 (2.2, 4.8) | 3.3 (2.2, 4.8) | 3.3 (2.1, 4.7) | 3.2 (2.1, 4.6) | 3.2 (2, 4.6) | 3.1 (2, 4.5) | 3 (1.9, 4.4) | 3 (1.9, 4.3) | 2.9 (1.8, 4.3) |
| _m<sup>1.7</sup> | 2.4 (1.6, 3.6) | 2.4 (1.6, 3.5) | 2.4 (1.6, 3.5) | 2.4 (1.5, 3.5) | 2.3 (1.5, 3.4) | 2.3 (1.5, 3.4) | 2.3 (1.4, 3.3) | 2.3 (1.4, 3.3) | 2.2 (1.4, 3.2) | 2.1 (1.3, 3.2) |
| _m | 3.7 (2.5, 5.4) | 3.7 (2.4, 5.4) | 3.7 (2.4, 5.3) | 3.6 (2.3, 5.2) | 3.6 (2.3, 5.2) | 3.5 (2.2, 5.1) | 3.5 (2.2, 5) | 3.4 (2.1, 4.9) | 3.3 (2.1, 4.8) | 3.1 (2, 4.7) |
| Mass | 130 (99, 165) | 130 (98, 165) | 127 (98, 164) | 127 (97, 163) | 126 (97, 162) | 126 (97, 160) | 127 (97, 159) | 124 (96, 156) | 122 (94, 153) | 116 (92, 149) |
| _bsa | 66 (55, 79) | 66 (53, 79) | 64 (52, 79) | 63 (51, 78) | 63 (50, 78) | 63 (50, 77) | 63 (51, 77) | 62 (50, 77) | 63 (50, 76) | 60 (49, 76) |
| _m<sup>1.7</sup> | 47 (37, 59) | 47 (37, 59) | 46 (37, 58) | 46 (37, 58) | 46 (37, 58) | 46 (37, 58) | 47 (37, 57) | 46 (36, 57) | 46 (36, 57) | 44 (36, 57) |
| _m | 72 (56, 89) | 72 (56, 89) | 70 (56, 89) | 70 (55, 88) | 70 (55, 88) | 70 (55, 88) | 70 (55, 87) | 69 (55, 86) | 69 (54, 85) | 66 (54, 83) |
| Wall thickness | 7.2 (6.2, 8.2) | 7.3 (6.2, 8.3) | 7.2 (6.3, 8.4) | 7.3 (6.3, 8.5) | 7.4 (6.3, 8.5) | 7.4 (6.4, 8.6) | 7.5 (6.4, 8.7) | 7.5 (6.4, 8.8) | 7.6 (6.5, 8.9) | 7.5 (6.5, 8.9) |
| _bsa | 3.7 (3.2, 4.3) | 3.7 (3.2, 4.2) | 3.6 (3.1, 4.2) | 3.6 (3.1, 4.2) | 3.7 (3.1, 4.3) | 3.7 (3.2, 4.3) | 3.8 (3.2, 4.4) | 3.8 (3.3, 4.5) | 3.9 (3.3, 4.5) | 3.9 (3.4, 4.6) |
| _m<sup>1.7</sup> | 2.6 (2.3, 3.1) | 2.6 (2.3, 3.1) | 2.6 (2.3, 3.1) | 2.7 (2.3, 3.1) | 2.7 (2.3, 3.2) | 2.7 (2.3, 3.3) | 2.8 (2.3, 3.3) | 2.8 (2.4, 3.4) | 2.9 (2.4, 3.4) | 2.9 (2.4, 3.4) |
| _m | 4 (3.5, 4.6) | 4 (3.5, 4.6) | 4 (3.5, 4.6) | 4 (3.5, 4.7) | 4.1 (3.5, 4.8) | 4.1 (3.5, 4.8) | 4.2 (3.5, 4.9) | 4.2 (3.6, 4.9) | 4.3 (3.6, 5) | 4.3 (3.7, 5) |
| midseptal | 7.5 (6.2, 8.8) | 7.5 (6.3, 8.8) | 7.5 (6.3, 8.9) | 7.7 (6.4, 9.2) | 7.8 (6.5, 9.4) | 7.9 (6.6, 9.7) | 8.2 (6.6, 9.9) | 8.3 (6.7, 10.1) | 8.4 (6.8, 10.1) | 8.3 (6.9, 10.1) |
| RV |  |  |  |  |  |  |  |  |  |  |
| EF | 53 (43, 64) | 54 (43, 64) | 54 (43, 64) | 54 (43, 65) | 54 (43, 65) | 54 (43, 65) | 55 (43, 66) | 55 (44, 66) | 55 (44, 67) | 56 (44, 67) |
| EDV | 194 (138, 258) | 194 (136, 257) | 193 (134, 254) | 191 (132, 252) | 188 (129, 249) | 185 (127, 245) | 182 (125, 242) | 177 (122, 238) | 172 (120, 233) | 166 (117, 229) |
| _bsa | 99 (73, 126) | 98 (71, 124) | 97 (70, 123) | 95 (68, 122) | 94 (67, 121) | 92 (65, 120) | 91 (64, 119) | 89 (62, 117) | 88 (61, 116) | 86 (59, 115) |
| _m<sup>1.7</sup> | 71 (52, 91) | 71 (51, 91) | 70 (51, 90) | 70 (50, 89) | 69 (49, 88) | 68 (48, 88) | 67 (48, 87) | 66 (47, 86) | 65 (46, 85) | 63 (45, 85) |
| _m | 107 (78, 139) | 107 (77, 138) | 107 (76, 137) | 105 (75, 136) | 104 (74, 135) | 102 (72, 133) | 101 (71, 132) | 99 (70, 130) | 97 (68, 129) | 94 (66, 127) |
| ESV | 91 (57, 124) | 90 (56, 125) | 89 (55, 125) | 88 (54, 124) | 86 (53, 123) | 84 (52, 121) | 82 (52, 118) | 79 (51, 114) | 76 (50, 110) | 73 (49, 106) |
| _bsa | 46 (30, 62) | 46 (30, 62) | 45 (29, 61) | 44 (28, 60) | 43 (28, 60) | 42 (27, 59) | 41 (27, 58) | 40 (26, 57) | 39 (26, 56) | 38 (25, 55) |
| _m<sup>1.7</sup> | 33 (21, 44) | 33 (21, 44) | 33 (21, 45) | 32 (21, 44) | 32 (20, 44) | 31 (20, 43) | 30 (20, 43) | 30 (19, 42) | 29 (19, 40) | 28 (19, 39) |
| _m | 50 (32, 67) | 50 (32, 68) | 49 (31, 68) | 49 (31, 68) | 48 (30, 67) | 47 (30, 66) | 46 (29, 64) | 44 (29, 62) | 43 (28, 60) | 41 (28, 58) |
| SV | 105 (70, 144) | 104 (71, 143) | 103 (71, 142) | 102 (69, 141) | 102 (67, 140) | 100 (65, 139) | 99 (64, 139) | 98 (61, 138) | 97 (58, 137) | 95 (53, 136) |
| _bsa | 53 (36, 71) | 52 (36, 70) | 52 (36, 70) | 51 (35, 70) | 51 (34, 69) | 50 (34, 69) | 50 (33, 69) | 49 (32, 69) | 49 (31, 69) | 48 (30, 69) |
| _m<sup>1.7</sup> | 38 (26, 51) | 38 (26, 51) | 38 (26, 51) | 37 (26, 51) | 37 (25, 51) | 37 (25, 51) | 37 (24, 50) | 36 (24, 50) | 36 (23, 50) | 36 (21, 50) |
| _m | 58 (39, 78) | 57 (40, 77) | 57 (40, 77) | 57 (39, 77) | 56 (38, 77) | 56 (37, 76) | 55 (36, 76) | 55 (35, 76) | 54 (34, 76) | 54 (31, 75) |
| CO | 6.6 (4.2, 9.9) | 6.6 (4.1, 9.8) | 6.5 (4.1, 9.7) | 6.5 (4, 9.7) | 6.4 (3.9, 9.6) | 6.3 (3.8, 9.5) | 6.3 (3.8, 9.4) | 6.2 (3.7, 9.3) | 6.1 (3.6, 9.2) | 6 (3.5, 9.2) |
| _bsa | 3.3 (2.2, 4.8) | 3.3 (2.1, 4.8) | 3.3 (2.1, 4.8) | 3.2 (2, 4.8) | 3.2 (2, 4.7) | 3.2 (2, 4.7) | 3.1 (1.9, 4.7) | 3.1 (1.9, 4.6) | 3.1 (1.8, 4.6) | 3 (1.8, 4.6) |
| _m<sup>1.7</sup> | 2.4 (1.5, 3.5) | 2.4 (1.5, 3.5) | 2.4 (1.5, 3.5) | 2.4 (1.5, 3.5) | 2.3 (1.5, 3.5) | 2.3 (1.4, 3.5) | 2.3 (1.4, 3.4) | 2.3 (1.4, 3.4) | 2.3 (1.4, 3.4) | 2.3 (1.3, 3.4) |
| _m | 3.6 (2.3, 5.4) | 3.6 (2.3, 5.3) | 3.6 (2.3, 5.3) | 3.6 (2.2, 5.3) | 3.5 (2.2, 5.3) | 3.5 (2.2, 5.2) | 3.5 (2.1, 5.2) | 3.5 (2.1, 5.2) | 3.4 (2, 5.2) | 3.4 (2, 5.1) |

Values are fitted mean (5th quantile, 95th quantile) from an additive model with CMR metric as an outcome and a non-linear smoothing function of continuous age, given by a restricted cubic spline basis and up to 20 knots, averaged by 5-year age group. CMR metrics were indexed to body surface area according to DuBois formula (\_bsa), body height in meters to the power of 1.7 (\_m<sup>1.7</sup>), and body height in meters (\_m). LV: left ventricle; RV: right ventricle; EF: ejection fraction; EDV: end-diastolic volume; ESV: end-systolic volume; SV: stroke volume; CO: cardiac output; BSA: body surface area.

### Supplementary Figure

**Supplementary Figure 1:**
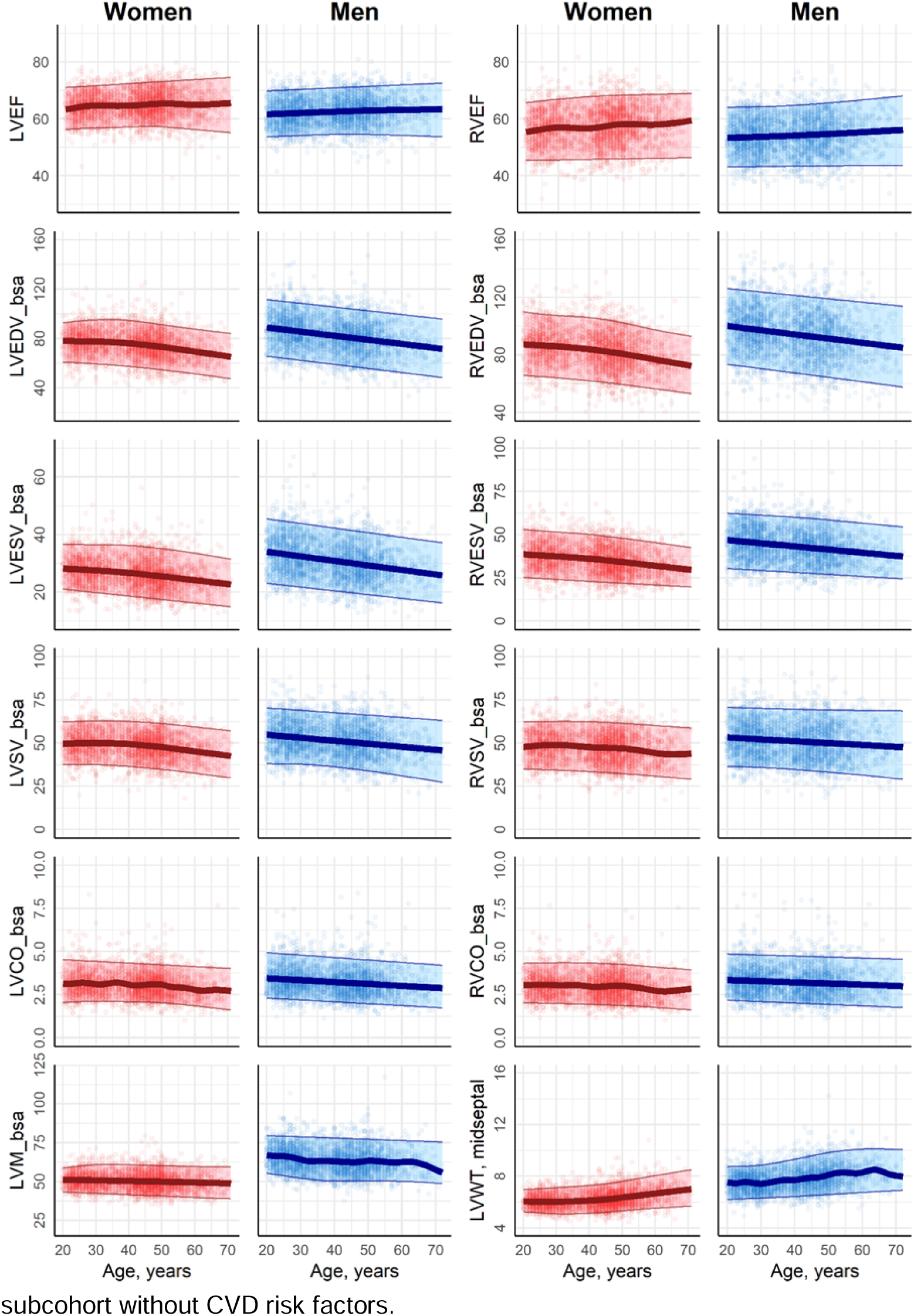
CMR reference ranges according to continuous age, derived from the healthy subcohort without CVD risk factors. Figures show fitted mean (5th quantile, 95th quantile) of the CMR metric on the y-axis, and continuous age on the x-axis for women (red) and men (blue). LV: left ventricle; RV: right ventricle; EF: ejection fraction; EDV: end-diastolic volume; ESV: end-systolic volume; SV: stroke volume; CO: cardiac output; LVM: Left ventricular mass; WT: wall thickness; _bsa: indexed to body surface area. EF is given in %. EDV_bsa, ESV_bsa, and SV_bsa are given in ml/m^2^. LVCO_bsa is given in l/min/m^2^. LVM_bsa is given in g/m^2^. LVWT is given in mm.

**Supplementary Figure 2:**
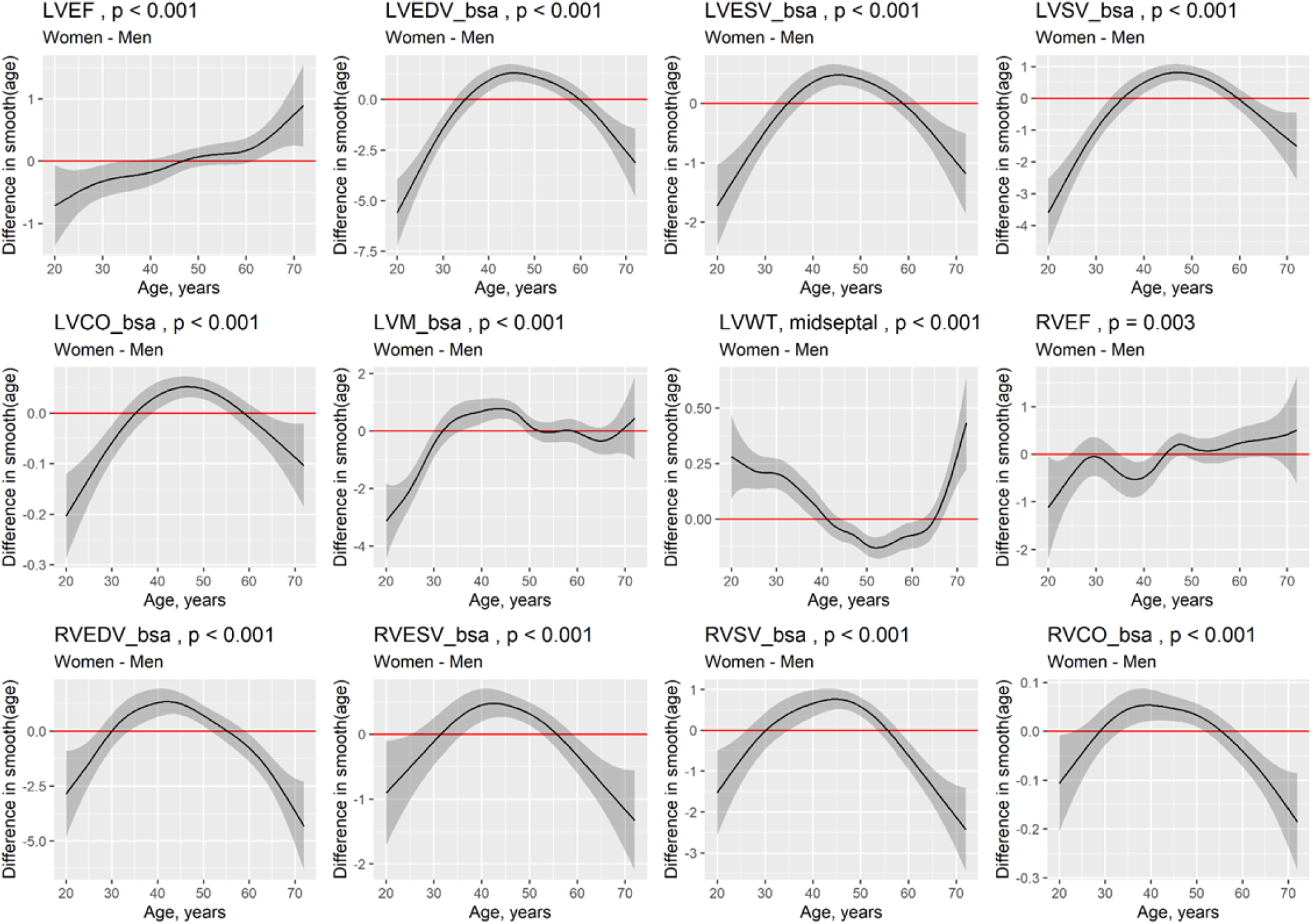
Difference of age effects between women and men Figure shows difference in smooth effect of age on CMR metric from generalized additive model between women and men (y-axis) for continuous age (x-axis) with 95% confidence interval (shaded area). Red horizontal line at 0 indicates where there is no difference in age effect between women and men. Positive values indicate stronger effects of age on CMR metric for women, whereas negative values indicate stronger effects of age on CMR metric for men. Plot created with R function gratia::difference_smooths.

